# The Effects of Social Isolation, Loneliness, and Nicotine Product Use: A Systematic Review and Meta-Analysis Evidence Update

**DOI:** 10.64898/2026.08.07.26359989

**Authors:** Rachael Pascoe, Christina Saliba, Anasua Kundu, Sharia Hoque, Andrew Milroy, Robert Schwartz, Michael Chaiton

## Abstract

**Background:** Loneliness and social isolation may contribute to tobacco and nicotine use, but existing studies have been inconsistent. This systematic review and meta-analysis examined these associations among adults amid the evolving nicotine product landscape.

**Methods:** A systematic search of PubMed, MEDLINE, PsycINFO, and CINAHL identified peer-reviewed quantitative studies published between 2014 and 2025 searched in February-April 2026. Eligible studies included adults aged ≥18 years examining loneliness and/or social isolation in relation to smoking or nicotine use. Two reviewers independently screened studies, extracted data, and assessed risk of bias (using the National Heart, Lung, and Blood Institute risk of bias tool). Random effects meta-analyses were conducted to estimate pooled odds ratio (OR) with 95% confidence intervals (CIs).

**Results:** 22 studies involving 273,954 participants met inclusion criteria, and 14 studies were included in the meta-analysis. A majority of the studies had low risk of bias. Meta-analysis findings showed that social isolation or loneliness was associated with significantly higher odds of nicotine product use (OR 1.84, 95% CI 1.48-2.29). Although no statistically significant association of nicotine product use and social isolation or loneliness (OR 1.35, 95% CI 0.72-2.53), some studies suggested bidirectional relationships, with smoking contributing to reduced social support and greater isolation over time. Sensitivity analysis showed the robustness of the meta-analysis findings. Subgroup analysis found no statistically significant differences were seen between subgroups defined by type of nicotine product, age groups, social isolation vs loneliness, measures of nicotine use behaviours and pre- vs post-COVID-19 pandemic period in the meta-regression. Limitations of included studies and analysis are discussed.

**Conclusions:** Loneliness and social isolation are significant psychosocial correlates of nicotine product use. Cessation interventions may benefit from integrating social support and mental health strategies alongside traditional nicotine dependence treatment.

## Introduction

The burden of tobacco and nicotine use remains substantial and represents a critical public health concern. Tobacco use is the leading preventable cause of premature death, disease, and disability in Canada accounting for an estimated 45,000 to 48,000 deaths annually (1). Within this context, smoking is increasingly recognized as a behavior shaped by social and structural determinants, including social isolation and loneliness that are related but distinct dimensions of social experience. Social isolation is an objective state characterized by limited social contact, small network size, or infrequent social participation, such as living alone or lacking close relationships (2). In contrast, loneliness is a subjective experience, reflecting a perceived gap between desired and actual social relationships (2). Both social isolation and loneliness are associated with adverse mental and physical health outcomes, including depression, cardiovascular disease, impaired sleep, and weakened immune functioning (3, 4). However, these constructs may differentially influence health and wellbeing outcomes as social isolation is associated with a higher relative risk of smoking (RR = 1.28) than loneliness (RR = 0.99) (5).

A growing body of literature has examined the relationship between loneliness, social isolation, and smoking (6–8). A longitudinal study of older adults in England found that smoking was associated with reduced social contact, a higher likelihood of living alone, and increased loneliness over time (9). Similarly, research among young adults indicates that smokers report greater loneliness, more anxious attachment styles, and poorer psychological well-being than non-smokers (10). Among students, loneliness has also been more commonly reported among smokers, suggesting that smoking may function as a coping mechanism for perceived social disconnection (11–13). However, some studies report non-significant associations with smoking behaviors (14, 15).

A systematic review published in 2015 examining loneliness and smoking found that only half of the included studies reported statistically significant associations (16). This variability underscores the need for more longitudinal, methodologically rigorous studies that examine potential mediators (e.g., stress, coping, social support) and moderators (e.g., age, socioeconomic status) to better clarify these relationships. Since then, the nicotine product landscape has also evolved with the emergence of electronic cigarettes (e-cigarettes), cigars, smokeless tobacco, and nicotine pouches, expanding how nicotine use is understood in relation to social contexts and social isolation or connectedness. Given these changes, an updated and comprehensive review is warranted. The aim of this study was to conduct a systematic review and meta-analysis to examine the association between loneliness, social isolation, and nicotine product use among adults aged 18 years and older. Specifically, the study objectives were to: (1) synthesize international evidence across all major nicotine product categories, (2) quantify the strength of associations where possible, and (3) explore variation in associations by product type, population characteristics, study design, and the impact of the COVID-19 pandemic.

## Methods

This systematic review and meta-analysis were prospectively registered on PROSPERO (CRD420261299611), and the Preferred Reporting Items for Systematic Reviews and Meta-Analyses (PRISMA) guideline was followed in reporting findings (17). No protocol was published and no amendments to information were made to the PRISMA registration.

### Search Strategy

A comprehensive literature search was performed from February to April 2026 across four electronic databases: PubMed, MEDLINE, PsycINFO, and CINAHL. The search strategy combined MeSH terms and equivalent subject headings with free-text keywords and truncated terms related to loneliness, social isolation, and smoking-related behaviors. The keyword strategy included: (lone* OR social isolat* OR isolat*) AND (smok* OR cig* OR tobacco* OR nicotine* OR vape* OR vaping* OR “nicotine pouch*”). MeSH terms were used to capture studies relating to all tobacco and nicotine products, including cigarettes, cigars, cigarillos, pipes, chewing tobacco or snuff, nicotine patches, tobacco-free nicotine pouches, snus, waterpipe or hookah, heated tobacco, e-cigarettes or vaping devices. Keywords were permitted to appear in any field to maximize search sensitivity. See Supplementary Figure 1 for the MEDLINE search strategy and Supplementary Figure 2 for the PRISMA reporting checklist.

### Study Selection

All identified records were imported into Covidence for screening and data management.

All stages were conducted independently by two reviewers, with disagreements resolved by consensus with a third reviewer.

### Eligibility Criteria

Studies were included if they examined adults aged 18 years and older from clinical or general population samples who used any nicotine product, including cigarettes, cigars or cigarillos, pipes, chewing tobacco or snuff, nicotine replacement therapy (e.g., patches), tobacco-free nicotine pouches (e.g., Velo, Zyn), snus, waterpipe or hookah, heated tobacco products (e.g., IQOS), or e-cigarettes. Studies focusing on adolescents under the age of 18 or exclusively non-smokers were excluded. The exposure of interest was loneliness and/or social isolation to maximize the sensitivity and to ensure comprehensive identification of relevant studies; studies that did not assess these constructs were excluded. Non-English publications, grey literature, and non-human studies were also excluded. Outcomes of interest included nicotine use patterns, such as initiation, frequency, or changes in use, in relation to loneliness and/or social isolation.

Eligible studies were limited to peer-reviewed quantitative research published in English from January 2014 onwards and involving human participants, as studies published before this date were included in a previous systematic review by Dyal & Valente (16). Studies excluded at the full-text screening stage, along with reasons for exclusion, were tracked within Covidence but are not reported in this manuscript.

### Data extraction

Data extraction was conducted using Covidence and a standardized data extraction form in Microsoft Excel. Extracted data included study identification details (e.g., authors, country, and setting), methodological characteristics (e.g., study design and recruitment strategy), and population characteristics (e.g., sample size, inclusion and exclusion criteria, and sample characteristics). Information on exposure measurement was collected, including how loneliness, social isolation, or social support were operationalized (e.g., Likert scales or validated instruments such as the NIH Toolbox Emotional Support Scale (18)).

The primary outcomes of interest were smoking-related behaviors, defined as: (1) smoking/nicotine use status (categorized as current, former/ex, or never user, as defined by each study), (2) change in smoking or nicotine use over time (e.g., self-reported increase, decrease, or no change, often relative to a specific event such as COVID-19 lockdown), (3) quantity of use (e.g., number of cigarettes smoked per day), and (4) smoking cessation or quit-related outcomes (e.g., cessation status, intention to quit), specifically at the latest time point under study. Nicotine product type was also extracted for each outcome (e.g., cigarettes, e-cigarettes/vapes, or combinations thereof) since studies varied in whether they assessed cigarette smoking specifically or nicotine/tobacco use more broadly. Where a study reported results for more than one product type (e.g., cigarettes and e-cigarettes separately) or more than one outcome measure (e.g., both current status and change in use), all compatible results were extracted rather than selecting a single measure. In addition to exposure and outcome data, information was extracted on other study characteristics relevant to interpretation, including country/setting, study design, sample size and age characteristics, and an assessment of risk of bias using the National Heart, Lung, and Blood Institute Quality Assessment Tool alongside any identified financial or authorship connections to the tobacco industry (e.g., journal or funding ties).

Data extraction was performed independently by two reviewers and subsequently checked by a third reviewer to ensure consistency and accuracy across studies. Where information required for extraction was missing, ambiguous, or not reported in sufficient detail (e.g., no mean age specified, or unspecified smoking measurement timeframe), reviewers noted this explicitly in extraction forms. Study authors were not contacted for clarification; instead, the relevant data point was coded as “not reported,” and the study was retained for qualitative synthesis but excluded from quantitative analyses requiring that specific data point. Extracted data is presented in Table 1.

**Table 1.**
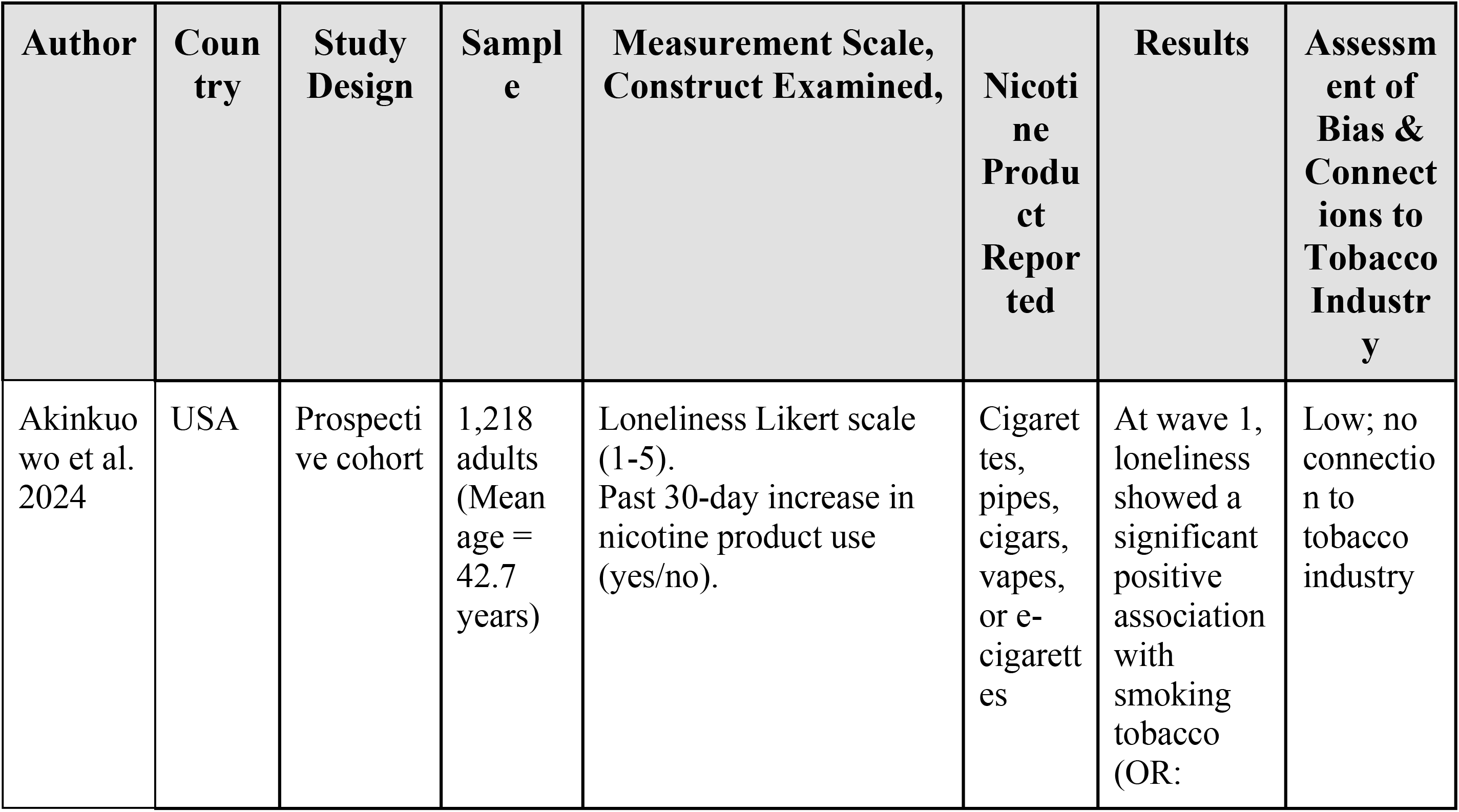
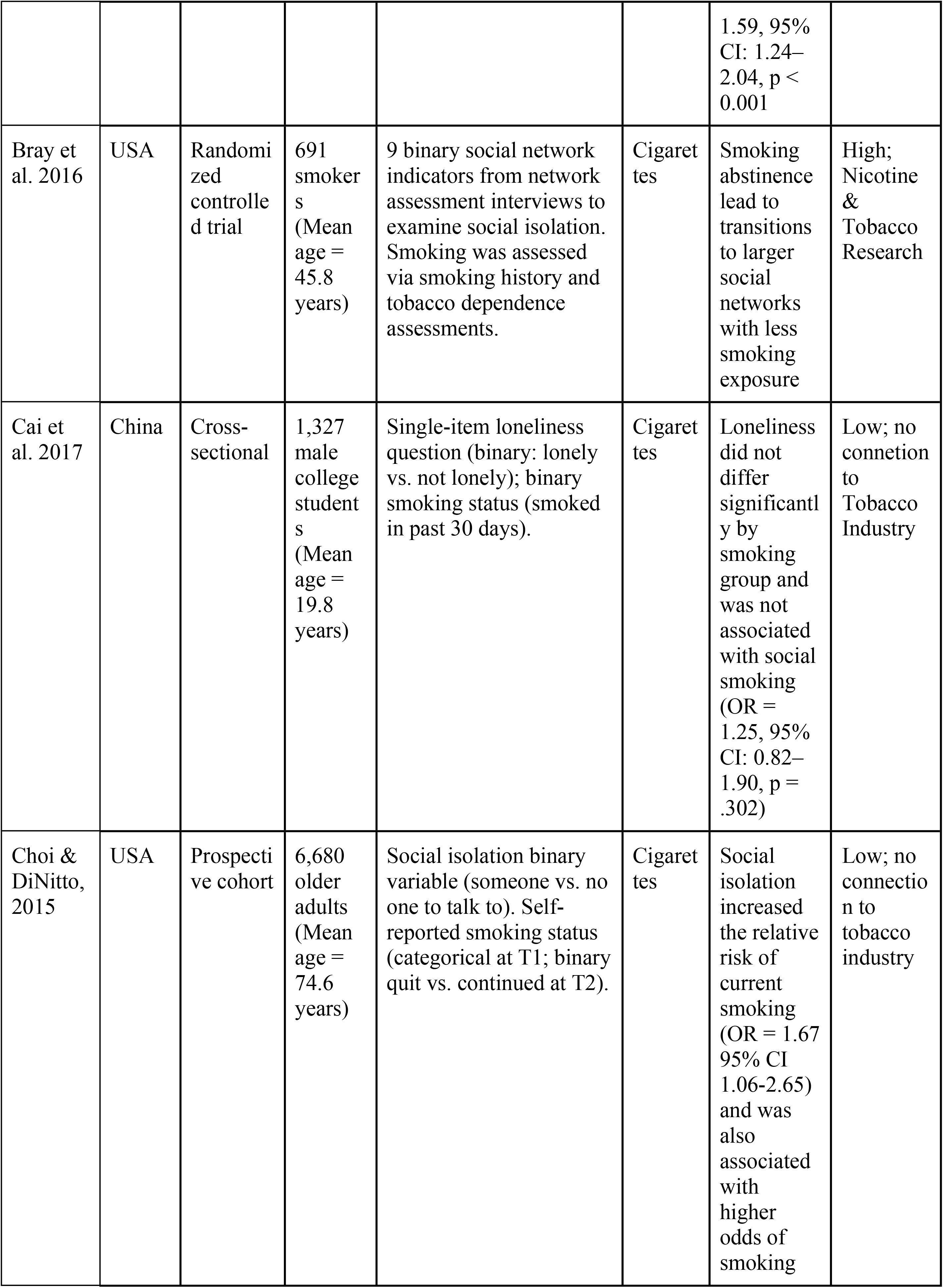
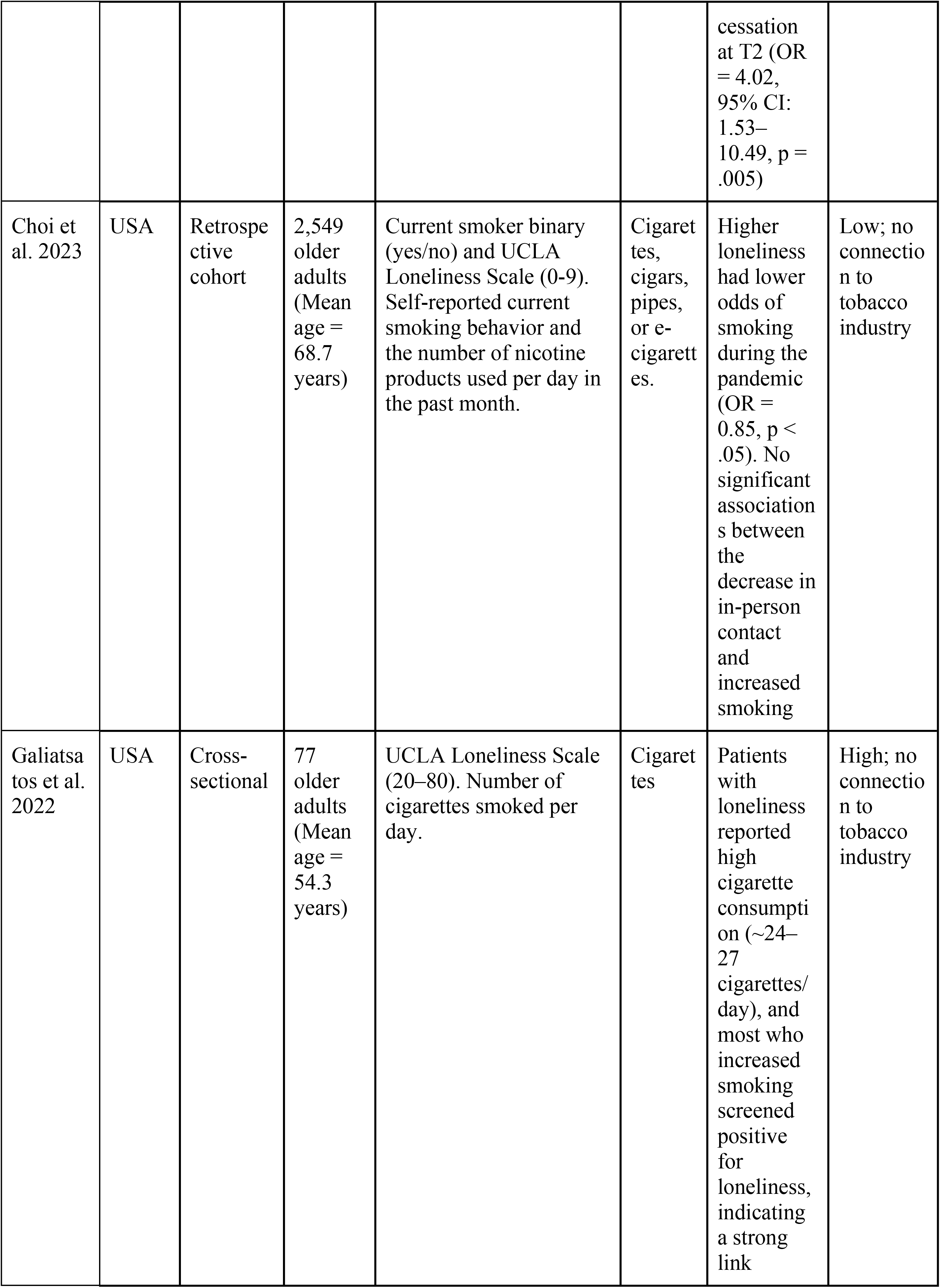
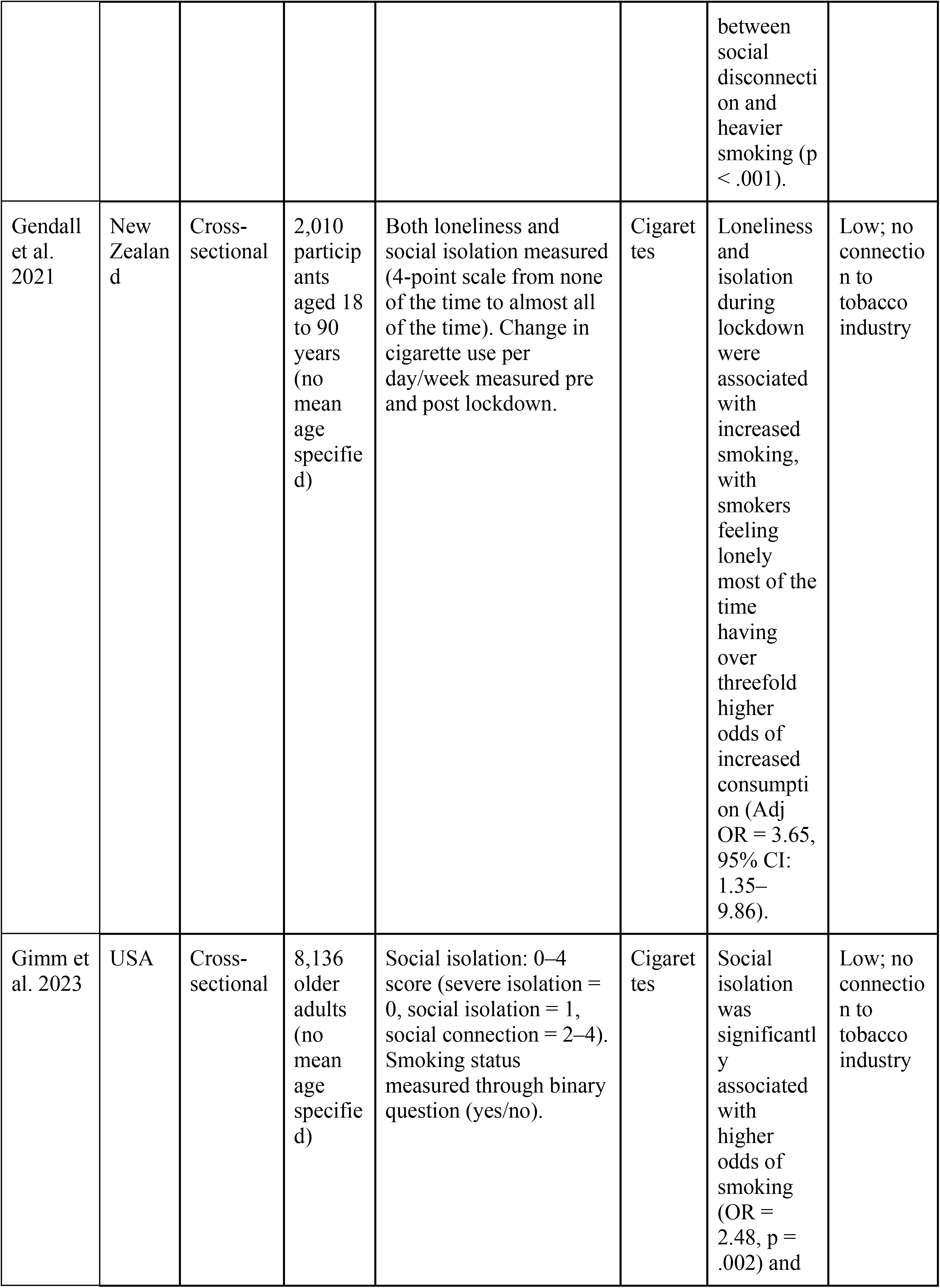
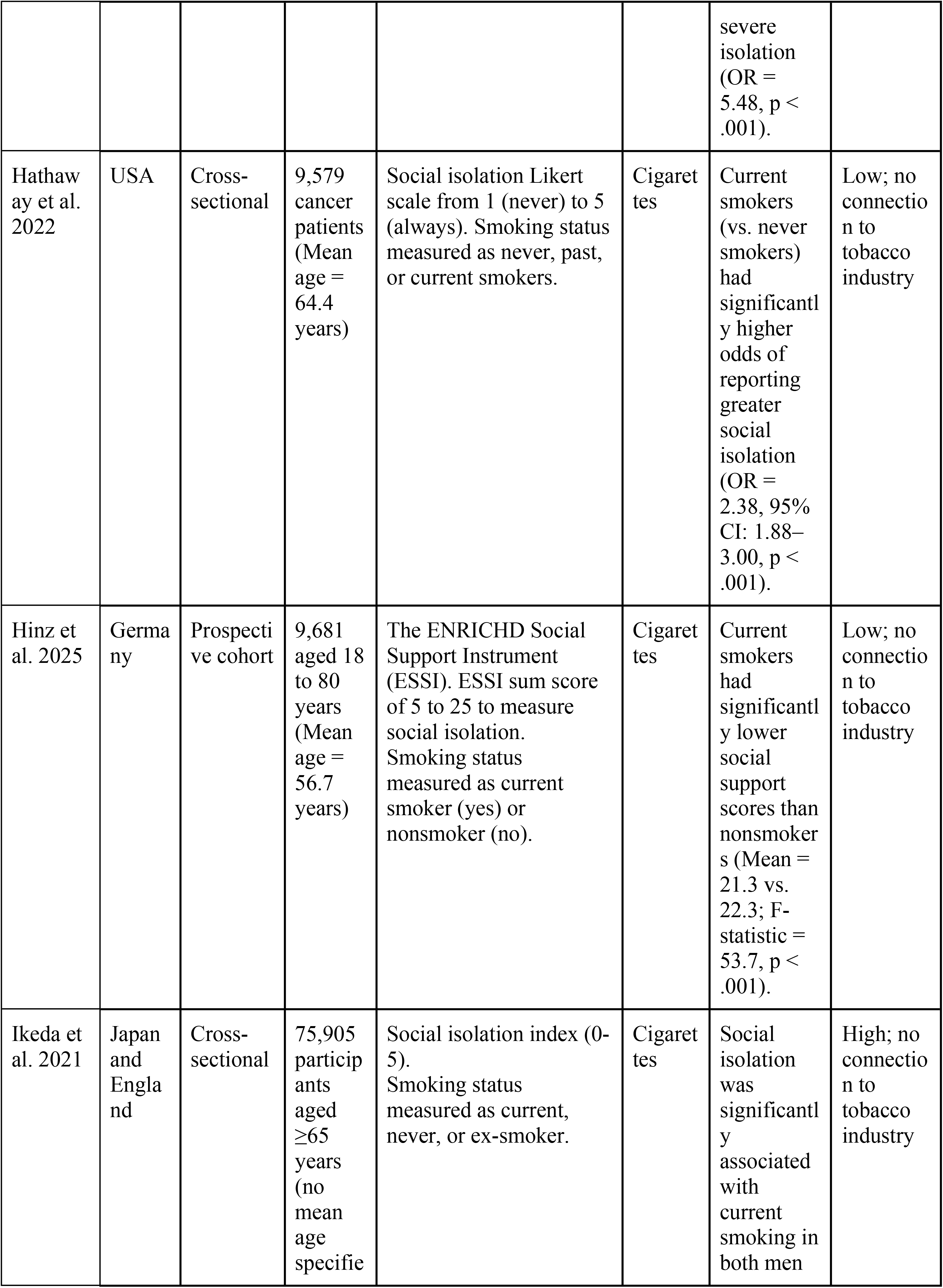
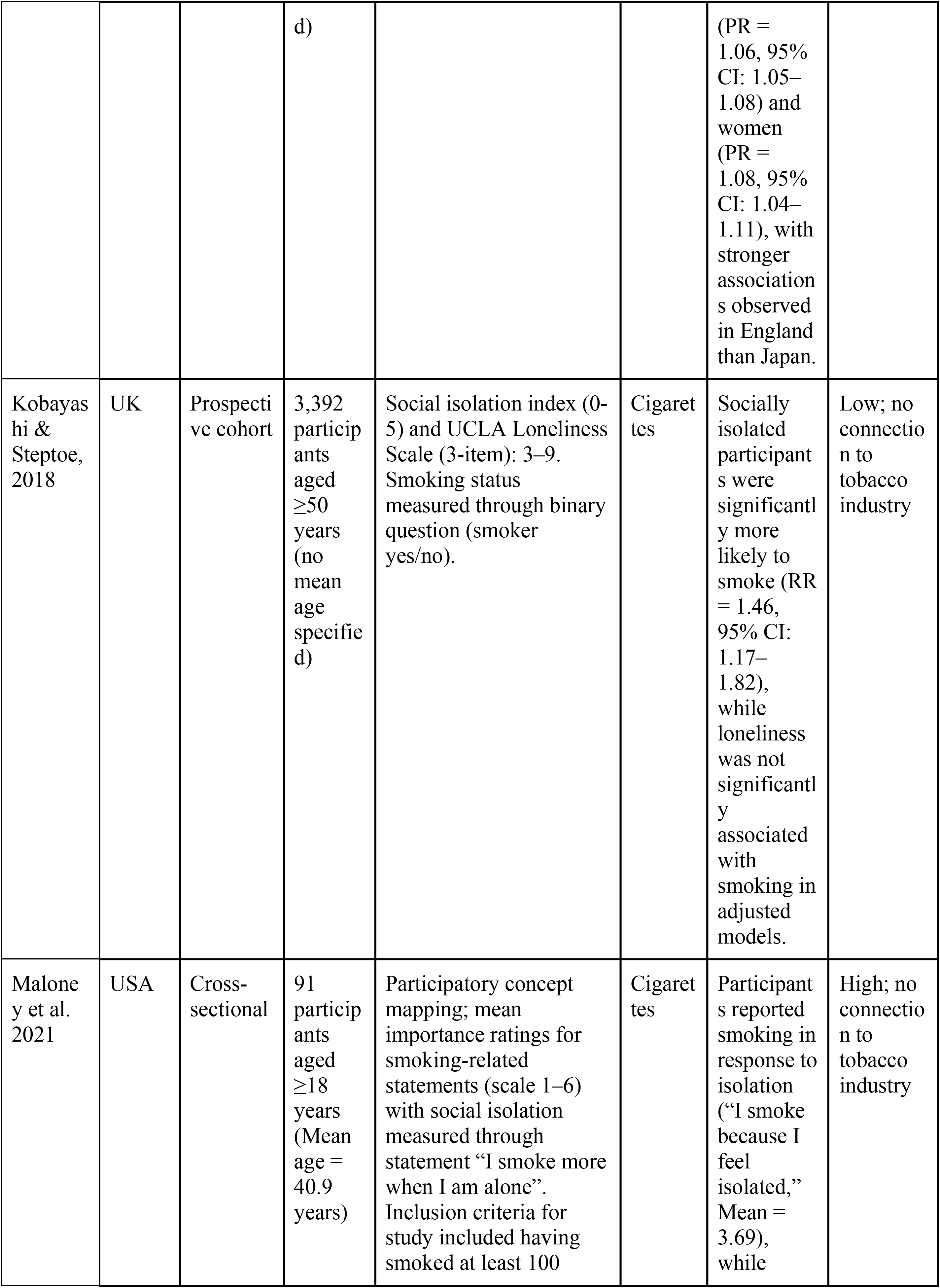
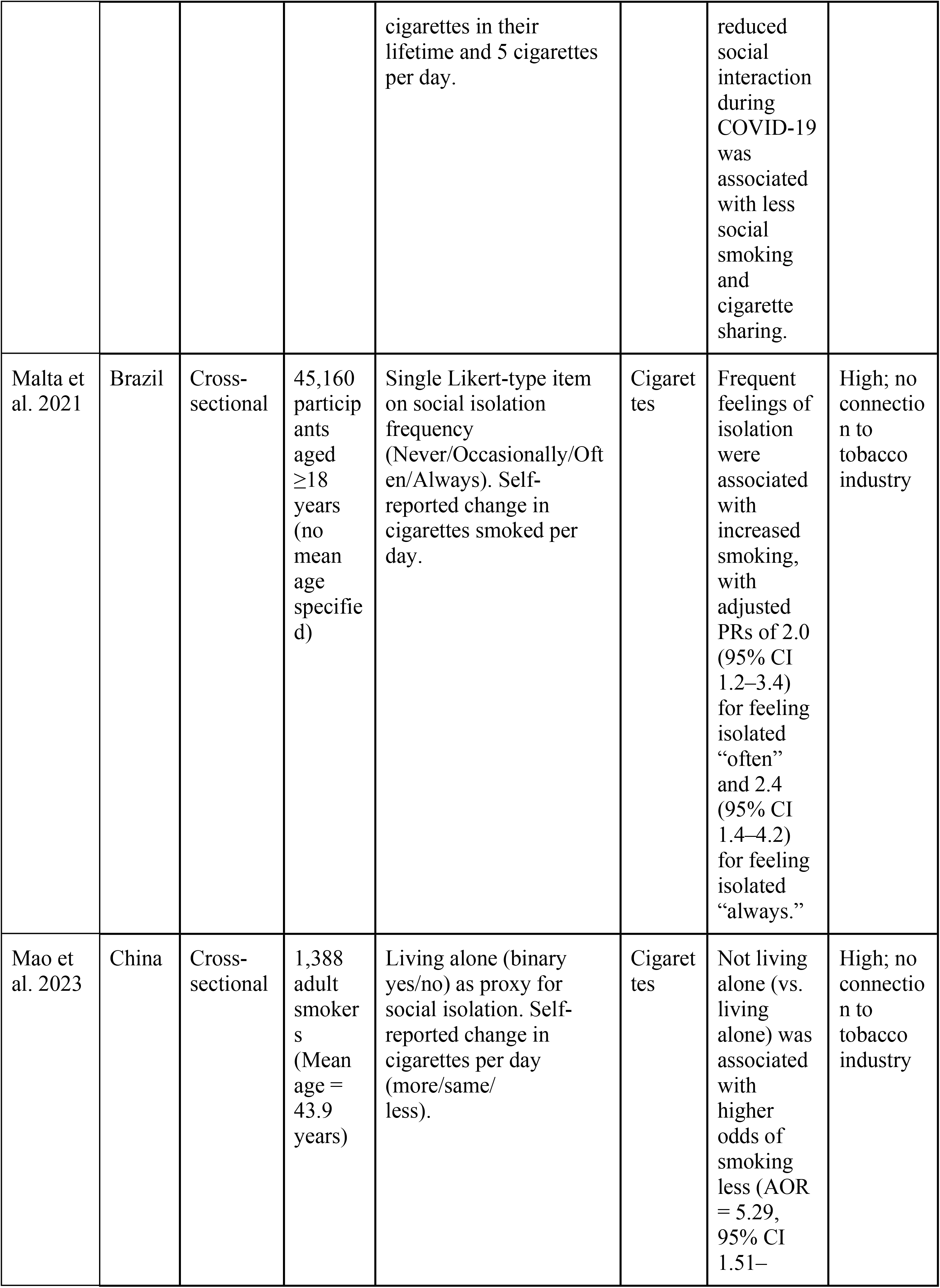
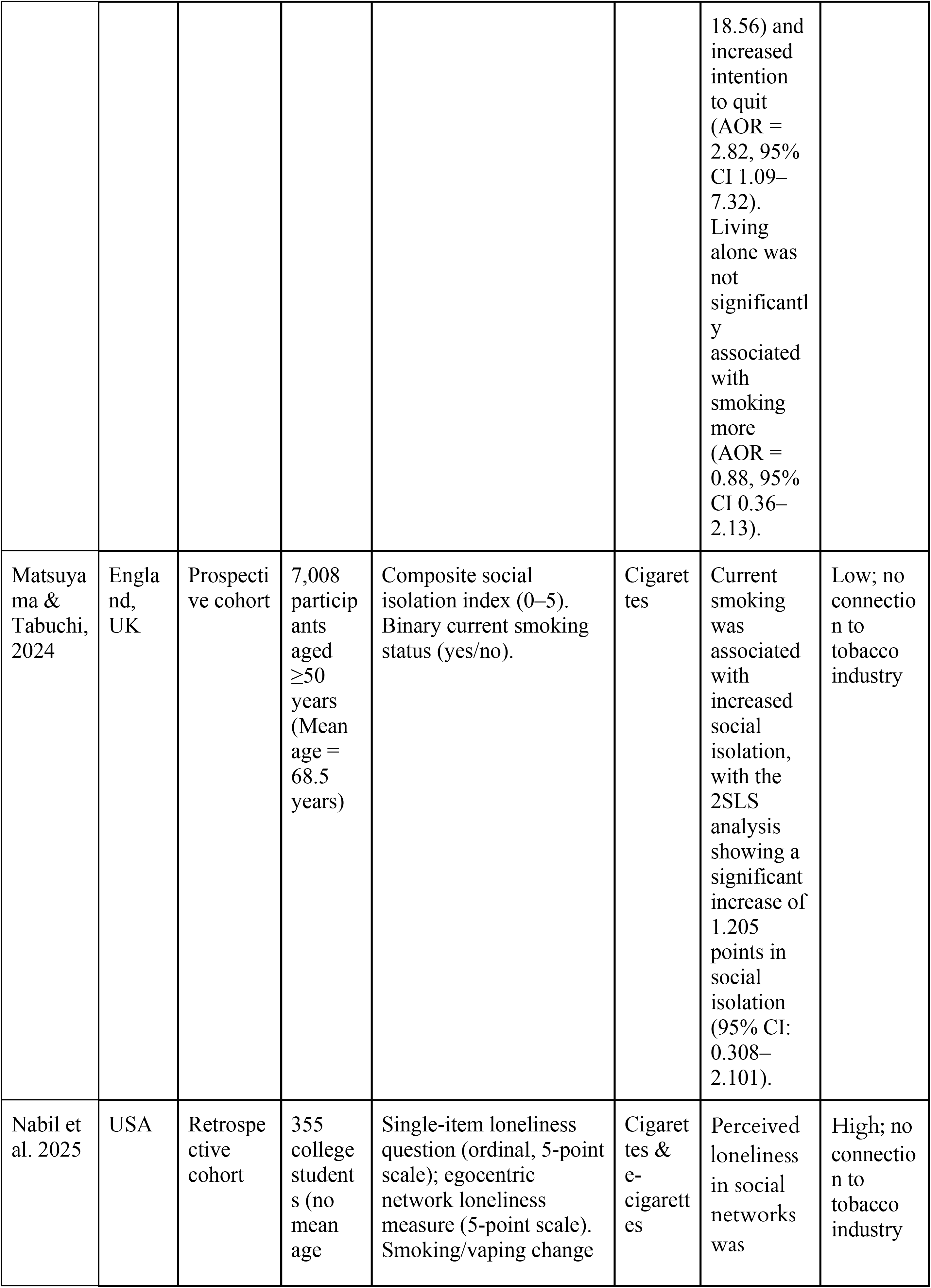
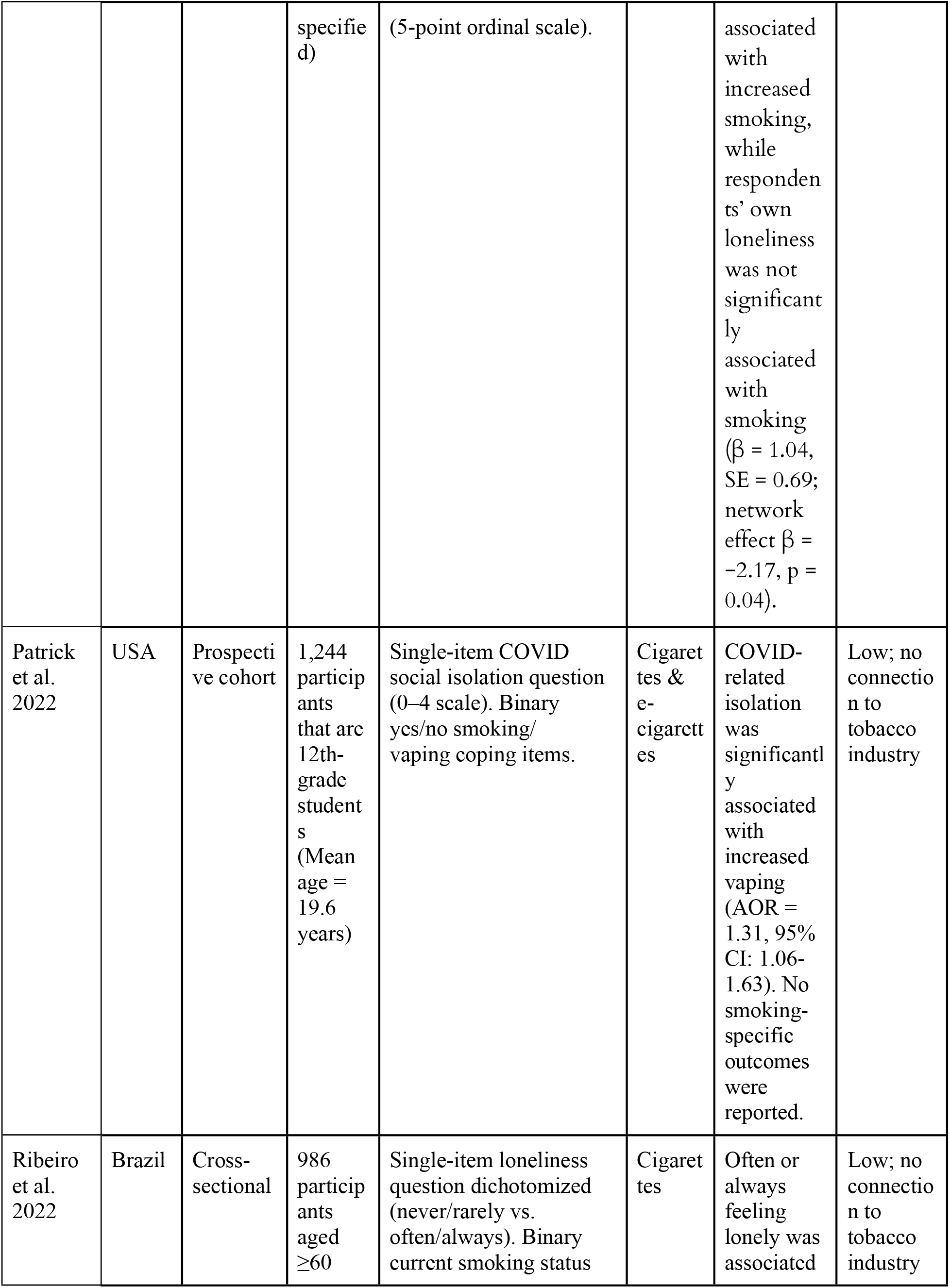
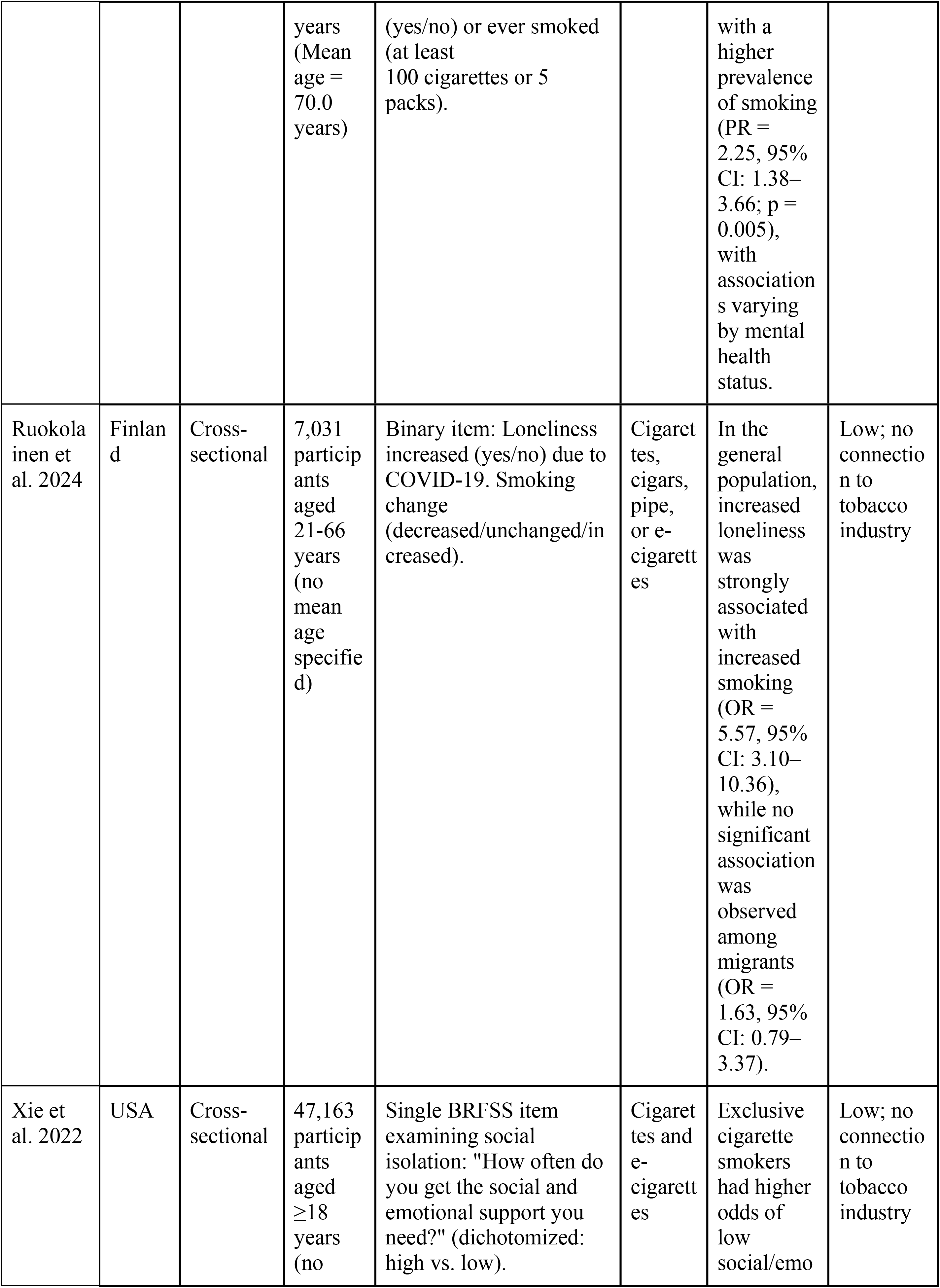
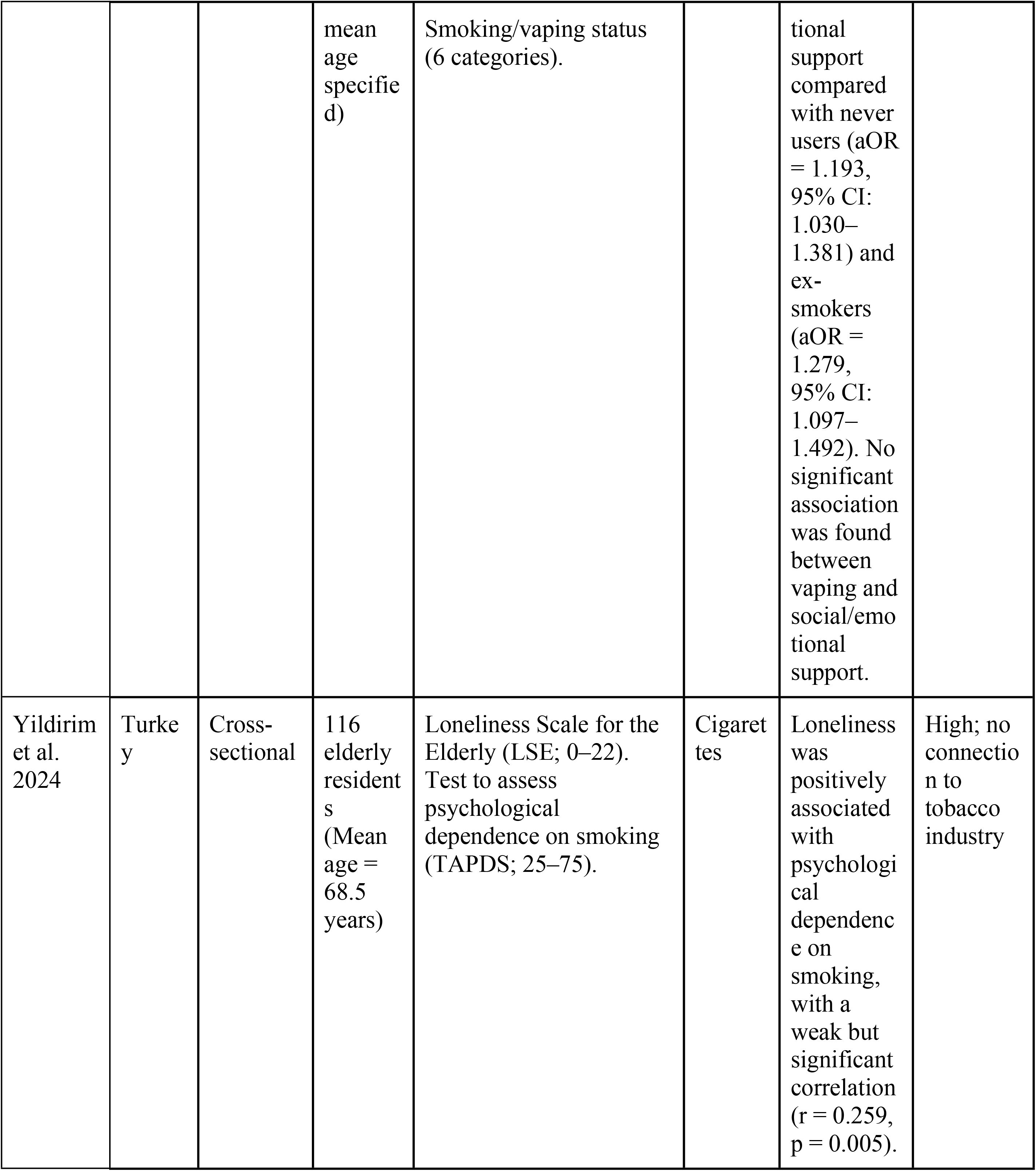
Summaries of studies included in the systematic review (N=22).

### Quality assessment

The methodological quality and risk of bias of included studies were assessed using the National Heart, Lung, and Blood Institute Study Quality Assessment Tool for Observational Cohort and Cross-Sectional Studies (NHLBI) (19). This tool evaluates domains including selection bias, exposure and outcome measurement, confounding, and attrition. Risk of bias assessments were conducted independently by two reviewers, with disagreements resolved with a third reviewer. Studies were not excluded based on quality assessment; however, the risk of bias was considered in interpreting the findings. A formal assessment of certainty of evidence (e.g., GRADE), was not conducted for the outcomes included in this meta-analysis. Instead, confidence in the pooled findings was considered narratively based on risk of bias ratings from the NHLBI tool, the magnitude and consistency of heterogeneity (I² statistics), and the precision of pooled estimates (width of 95% confidence intervals), alongside sensitivity analyses and assessment of publication bias where applicable.

### Data synthesis and meta-analysis

Meta-analysis was conducted to assess the impact of social isolation and loneliness on nicotine product use and vice versa. All statistical analysis was conducted using R statistical software version 4.5.3. Prior to synthesis, subgroup analyses were pre-specified according to the following characteristics: (1) nicotine product type (2) age of sample, (3) type of exposure measured (social isolation vs. loneliness vs. social support), (4) method of nicotine product use measurement, and (5) timing relative to the COVID-19 pandemic. Study characteristics were tabulated in a data extraction table and compared against subgroup categories. Studies were eligible for inclusion in the meta-analysis if they reported sufficient effect estimates (e.g. relative risk [RR], Odds Ratio [OR], or Incidence Rate Ratio [IRR]), and corresponding 95% confidence intervals under a rare events assumption. Studies that did not provide sufficient data on effect estimates, used incompatible outcome definitions, or exhibited substantial methodological heterogeneity were excluded from quantitative synthesis and summarized narratively. Reported ORs and RRs were converted to the logarithmic scale prior to pooling. Standard errors were derived from the reported 95% confidence intervals. Pooled effect estimates were calculated and presented as pooled ORs with 95% confidence intervals. Non-harmonized common scales were extracted and used in pooled analysis for exposure and outcome measures. Random-effects models were primarily used to account for between-study variability. Forest plots were created to summarize information from individual studies. Results were considered statistically significant in the meta-analysis when the 95% CI did not cross the null value. Heterogeneity was assessed primarily using the I^2^ statistic, with values of <25%, 25–50%, and >50% considered to indicate low, moderate, and high heterogeneity, respectively (20). Meta-regression analyses were conducted to examine whether subgroup characteristics moderated the pooled effect estimates and statistical significance.

Sensitivity analyses were performed using a leave-one-out approach, where studies were sequentially removed to assess robustness of pooled estimates. As the primary meta-analysis combined ORs and RRs as effect estimates, an additional sensitivity analysis was conducted by performing separate meta-analyses for studies reporting ORs and those reporting RRs to assess whether the pooled effect estimates differed by effect measure. Funnel plots were examined to assess potential publication bias. Asymmetry was further assessed using Egger’s test (21).

## Results

### Study selection

The literature search of academic databases retrieved 1,747 articles (Fig. 1). A total of 266 studies were excluded, as they were found to be duplicates. 1,481 studies were screened; 1,434 studies were excluded as they were unrelated to the topic of interest. Of the remaining 47 studies for full-text review, 25 studies were further excluded as they did not meet the predefined eligibility criteria (Fig 1). In total, 22 studies remained in this systematic review, and 14 articles were identified as suitable for the meta-analysis.

**Figure 1.**
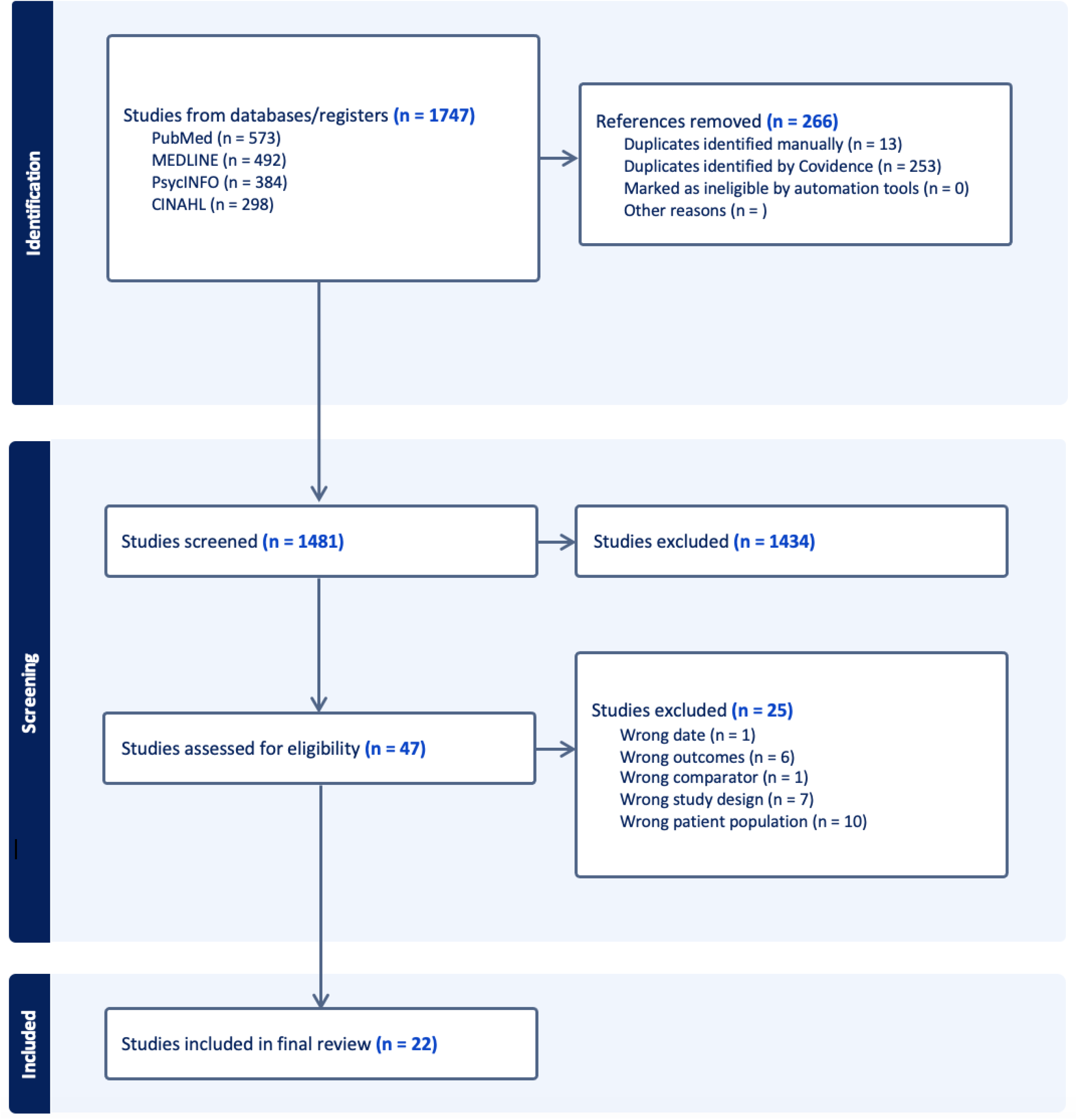
PRISMA flow diagram showing the study selection process.

### Study Characteristics

All studies included in the systematic review were published between 2015 and 2025 and had an overall total of 231,787 participants (Table 1). Of the 22 studies, ten studies were conducted in the United States (8, 22–31), two in Brazil (32, 33), two studies each in China (34, 35) and the United Kingdom (14, 36), and one study each in New Zealand (37), Germany (6), Finland (38), Turkey (39), Japan/England combined (40). The evidence base was methodologically diverse but predominantly observational, with cross-sectional designs being the most common (26–29, 32–35, 37–41) (n = 13), followed by prospective cohort studies (6, 14, 22, 24, 31, 36) (n = 6), retrospective cohort studies (25, 30) (n = 2), and one randomized controlled trial (23). All studies included adults aged 18 years and older, with most focusing on older populations (6, 14, 24, 40). Several studies specifically targeted older adults aged ≥50 or ≥60 years using large cohort datasets and population-based surveys (7, 24, 33, 36, 40). A narrative summary of study characteristics and risk of bias specific to each synthesis/subgroup was not reported; study design, sample characteristics, and risk-of-bias ratings for individual studies are presented in the data extraction table (Table 1).

### Quality assessment in studies

Of the 22 studies, 64% (n = 14) had a low risk of bias, 14% had a moderate risk of bias (n = 3), and 23% (n = 5) had a high risk of bias. None of the 22 studies had confirmed tobacco industry association or funding reported.

### Meta-analysis

Of the 22 studies extracted during the systematic review, 14 studies were found eligible for meta-analysis (8, 14, 22, 24, 27, 28, 31–35, 37, 38, 40). Eight studies were excluded, as they did not report effect sizes suitable for combining in the meta-analysis. Two meta-analyses were conducted. In the first meta-analysis, 13 studies were included that assessed the impact of social isolation or loneliness on nicotine product use. Eleven of these studies were assessed as having a low risk of bias (14, 22, 24, 27, 28, 31–35, 37, 38, 40), and two were assessed as having a high risk of bias (32, 40). Three included studies reported effect sizes for different sample groups and were therefore reported twice in the meta-analysis, resulting in a total of 16 effect estimates (Figure 2). Specifically, Akinkuowo et al. reported separate effect estimates for smoking and vaping, Ikeda et al(40) reported sex-specific estimates, and Ruokolainen et al. (38) reported separate estimates for Finnish migrants and the general population. The random effects meta-analysis demonstrated a statistically significant risk of nicotine product use among people facing social isolation or loneliness compared to people who did not experience either (OR 1.84; 95% CI 1.48-2.29; N=161,656). Heterogeneity was high (I²= 92.3%) between the studies (Figure 2).

**Figure 2.**
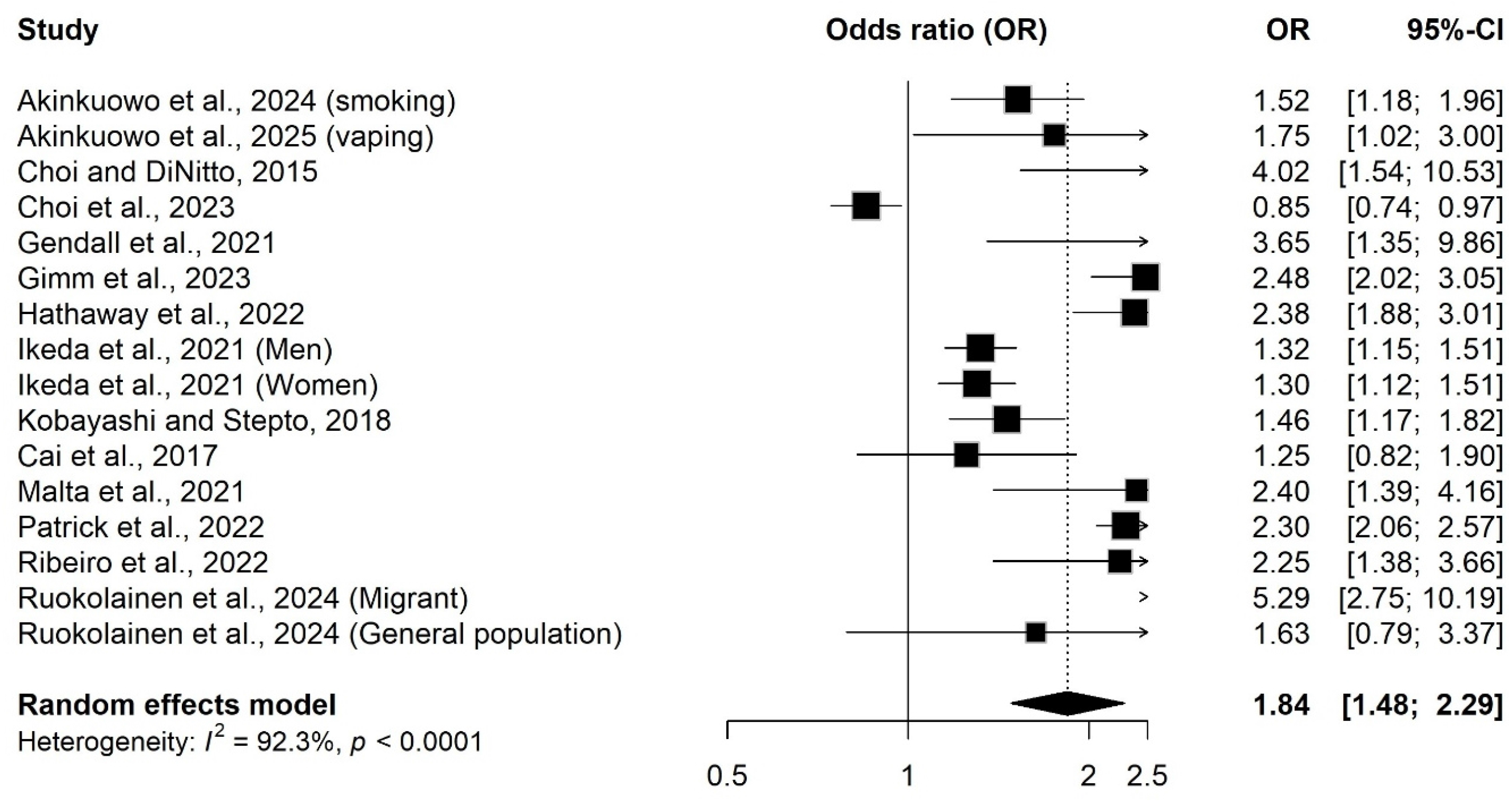
Forest plot showing effects of social isolation or loneliness on nicotine product use.

In the second meta-analysis, only two studies were included that assessed the impact of nicotine product use on social isolation or loneliness (8, 28). Both papers were rated as having low risk of bias. As Xie et al. (8) reported effect sizes for both vaping and smoking; both effect estimates were included in the meta-analysis, resulting in total 3 effect estimates (Figure 3). The random-effects meta-analysis found no statistically significant difference in risk of social isolation or loneliness among nicotine product users compared to non-users (OR = 1.35, 95% CI: 0.72–2.53). Between-study heterogeneity was substantial (I² = 92.9%) in this analysis (Figure 3).

**Figure 3.**
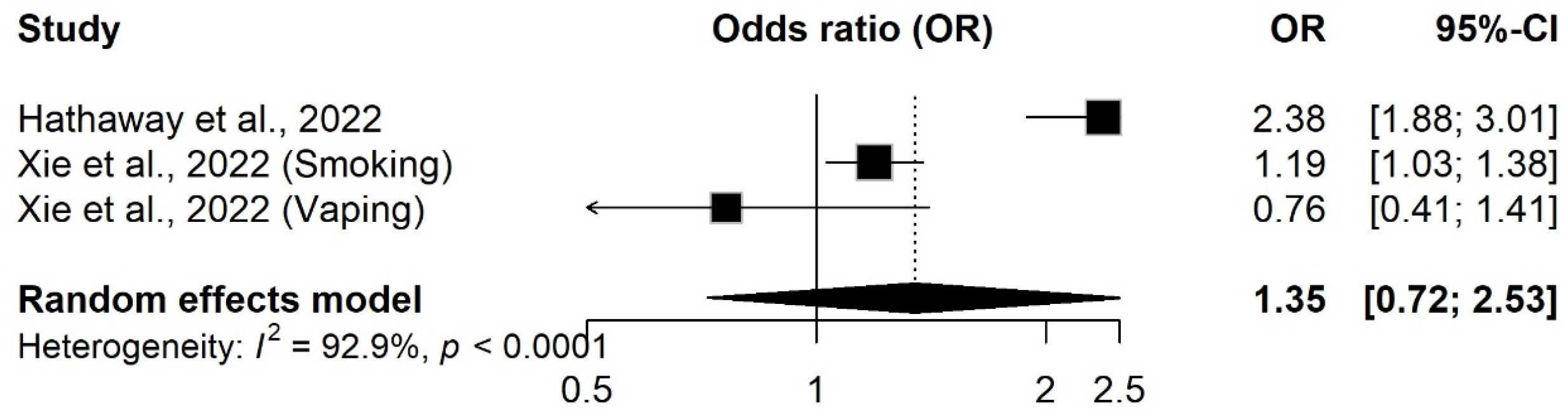
Forest plot for the impact of nicotine product use on social isolation or loneliness.

### Sensitivity analysis

Leave-one-out sensitivity analyses showed that omission of individual studies did not materially alter the overall pooled effect estimates of either meta-analysis. The pooled effect size for the first meta-analysis remained consistently statistically significant with ORs ranging between 1.73 and 1.94 and 95% CIs ranging from 1.42 to 2.39 (Table 2), indicating higher odds of nicotine product use in people experiencing social isolation or loneliness. Heterogeneity remained high across analyses (I^2^ =85.5%-92.8%), indicating that no single study was responsible for the substantial between-study variability observed in the meta-analysis. These results support the robustness of the observed association, although the consistently high heterogeneity suggests important differences in study populations, measures, or methodologies that may contribute to variation in effect sizes. The funnel plot showed no evidence of substantial asymmetry (Supplementary Figure 3) and Egger’s regression test was not statistically significant (t = 1.10, df = 14, p = 0.290), suggesting no evidence of small-study effects or publication bias.

**Table 2.**
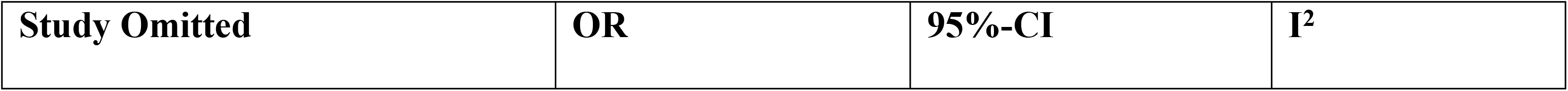
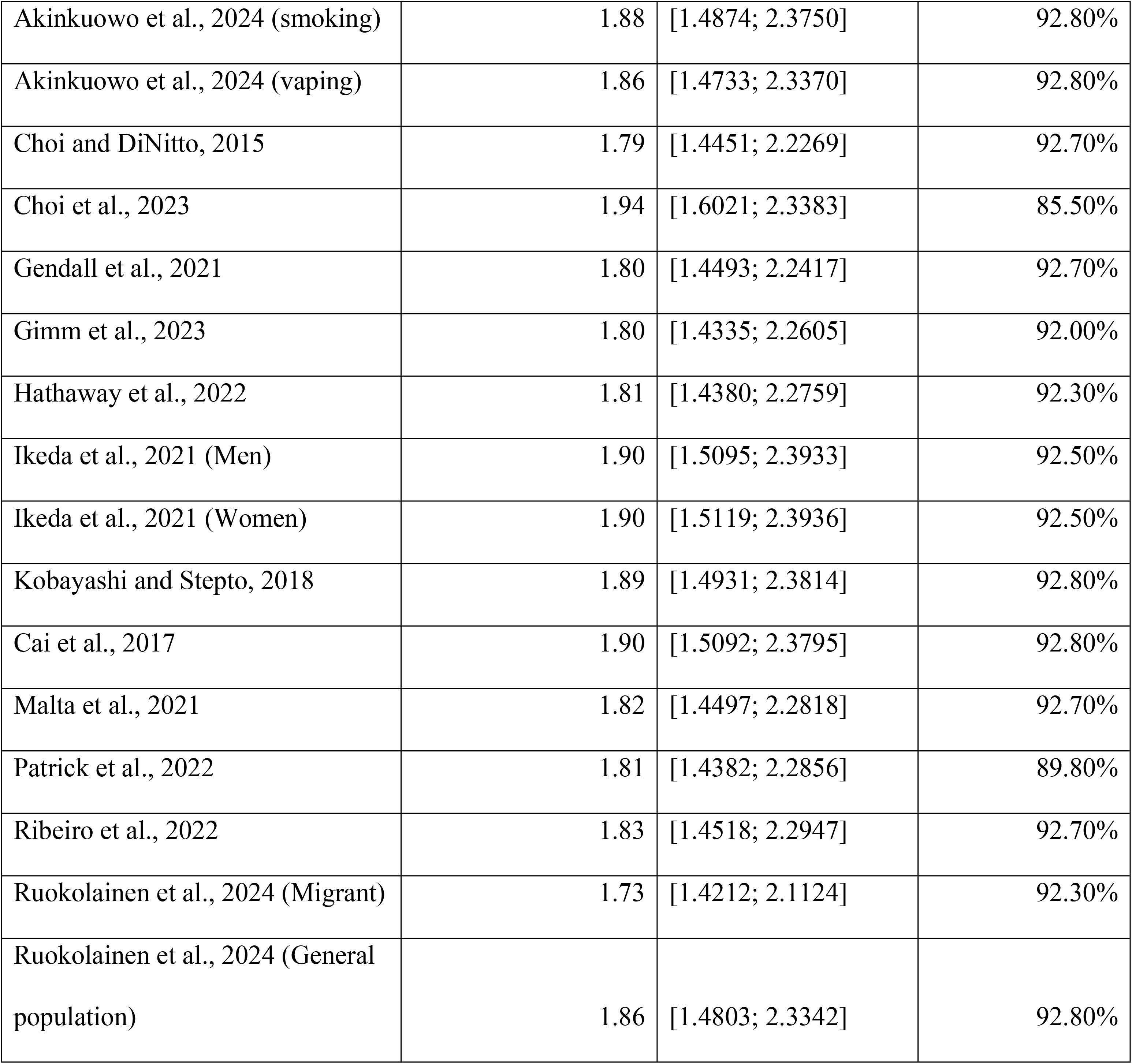
Leave-one-out sensitivity analysis of studies examining effects of social isolation and loneliness on nicotine product use (random effects model)

Removing each study individually from the second meta-analysis assessing the impact of nicotine product use on social isolation or loneliness resulted in statistically non-significant pooled ORs ranging between 1.06 and 1.67, and 95% CIs crossing the null value (Table 3), indicating robustness of the pooled estimate. A substantial reduction in heterogeneity (I^2^=48%) was observed from the removal of Hathaway et al.(28), suggesting its disproportionate contribution to between-study variability. As fewer than 10 studies were included in this meta-analysis, publication bias was not assessed (42).

**Table 3.** Sensitivity analysis of the meta-analysis of papers observing the impact of nicotine product use on social isolation and loneliness.

| Study Omitted | OR | 95% CI | I<sup>2</sup> |
| --- | --- | --- | --- |
| Omitting Hathaway et al., 2022 | 1.06 | [0.72; 1.56] | 48% |
| Omitting Xie et al., 2022 (Smoking) | 1.40 | [0.46; 4.26] | 91.30% |
| Omitting Xie et al., 2022 (Vaping) | 1.67 | [0.85; 3.29] | 95.80% |

Sensitivity analysis separating studies that reported effect estimates as ORs (14, 22, 24, 25, 27, 28, 37) and RRs (31–34, 38, 40) revealed consistently similar results. Social isolation or loneliness was associated with both higher odds of nicotine product use (OR 1.83; 95% CI 1.31–2.55; I² = 93.8%) (Supplementary Figure 4) and higher risk of nicotine product use (RR 1.88; 95% CI 1.39–2.53; I² = 90%) (Supplementary Figure 5). These findings indicate that combining ORs and RRs did not materially affect the overall pooled estimate or the direction of the association.

### Subgroup analysis

Subgroup analyses were conducted only for the first meta-analysis. As the second meta-analysis included only two studies, subgroup analyses were not performed for this analysis.

#### E-cigarette versus cigarette use

While the majority of the studies (14, 22, 24, 25, 27, 28, 32–34, 37, 38, 40) examined the impact of social isolation or loneliness on combustible cigarette smoking, only two studies (22, 31) examined the impact on e-cigarette use (Supplementary Fig 6). Overall, it was found that individuals with social isolation or loneliness had significantly higher odds of both smoking (OR 1.83; 95% CI 1.43-2.34; I^2^=90.5%) and vaping (OR 2.27; 95% CI 2.04-2.54; I^2^ = 0%) compared to people who did not experience either. Meta-regression analysis testing product type (e-cigarette vs cigarette) as a moderator was statistically non-significant (QM(df=1) = 0.14, *p* = 0.711), indicating no substantial subgroup differences (Supplementary Table 3).

#### Age groups

Subgroup analysis was conducted to compare the impact on nicotine product use among the following age groups: young adults (aged 18-25), middle-aged adults (ages 26-40), upper middle-aged adults (aged 41-60), and seniors (aged 61+ years). The target population in most of the studies were seniors (14, 25, 27, 28, 33, 40), while two studies were conducted among young adults (31, 34), two among middle aged adults (32, 38), and three among upper middle aged adults (22, 24, 37). Overall, social isolation or loneliness was associated with higher odds of nicotine product use among all age groups, but the effect among the young adults was statistically non-significant (OR 1.76; 95% CI 0.97-3.18; I^2^=88.8%) (Supplementary Figure 7). The highest statistically significant impact was seen among middle-aged adults (OR 2.76; 95% CI 1.44-5.33; I^2^=67%), followed by upper middle-aged adults (OR 2.07; 95% CI 1.33-3.23; I^2^= 50.8%), and seniors (OR 1.58, 95% CI 1.17-2.13, I^2^=94.4%). Meta-regression analysis testing age group as a moderator was statistically non-significant (QM(df=3) = 3.27, *p* = 0.352), indicating no substantial subgroup differences (Supplementary Table 3).

#### Social Isolation vs Loneliness

A subgroup analysis was conducted by exposure type, comparing studies that examined the impact of loneliness (22, 25, 33, 34, 38), social isolation (24, 27, 28, 31, 32, 40), and both social isolation and loneliness (14, 37) on nicotine product use (Supplementary Figure 8).

Overall, a significantly higher odds of nicotine product use among people experiencing only social isolation was found (OR 1.99; 95% CI 1.53-2.59; I^2^=92%) or loneliness (OR 1.69; 95% CI 1.12-2.54; I^2^=89.1%). Studies (n=2) examining both concepts demonstrated a higher, but statistically non-significant, effect size than either individually (OR 2.02; 95% CI 0.86-4.77; I^2^=92.2%). However, due to a low sample size (n=2), this result should be interpreted cautiously. Meta-regression analysis testing exposure type as a moderator was statistically non-significant (QM(df=2) = 0.70, *p* = 0.705), indicating no substantial subgroup differences (Supplementary Table 3).

#### Measures of Nicotine Product Use

Subgroup analysis was conducted to compare studies according to how nicotine product use was measured (Supplementary Figure 9). Some studies measured increases in reported nicotine product use over time (22, 25, 31, 32, 37, 38), while some studies measured current smoking status at the time of study (14, 24, 27, 28, 33, 34, 40). Overall, higher odds of association were observed between social isolation or loneliness and increased nicotine use (OR 1.97; 95% CI 1.33-2.92, I^2^=95.1%) and current use status (OR 1.76; 95% CI 1.38-2.23; I^2^=86.2%). However, meta-regression analysis testing measures of nicotine product use as a moderator was statistically non-significant (QM(df=2) = 0.11, *p* = 0.741), indicating no substantial subgroup differences (Supplementary Table 3).

#### Effects of COVID-19

A subgroup analysis of studies with data collected after COVID-19 public health policies (22, 25, 28, 31, 32, 37, 38) were compared with those conducted prior to COVID-19 (14, 24, 27, 33, 34, 40). Social isolation and loneliness measured after the COVID-19 pandemic demonstrated a stronger effect on nicotine product use (OR 2.01; 95% CI 1.43-2.83; I^2^= 94.8%) than studies collected before COVID-19 (OR 1.66; 95% CI 1.30-2.29; I^2^=83.9%) (Supplementary Figure 10). While accounting for consistently high heterogeneity, these findings may demonstrate that the social isolation caused by social distancing policies during the COVID-19 pandemic may have had a particularly strong effect on nicotine product use from loneliness/social isolation. However, meta-regression analysis testing COVID-19 as a moderator revealed statistically non-significant results (QM(df=1) = 0.44, *p* = 0.507), indicating no substantial subgroup differences (Supplementary Table 3).

### Narrative review findings

#### Social isolation, Loneliness and Nicotine Product Use

Among studies not included in the meta-analysis, four studies examined loneliness and smoking or nicotine product use (6, 8, 25, 26, 29, 30, 36, 39). Findings were more heterogeneous than those observed for social isolation. Four studies reported positive associations between loneliness and smoking behaviors [24,27,28,36]. These studies reported that greater loneliness was associated with increased nicotine product use. Participants experiencing higher levels of loneliness were more likely to report heavier cigarette consumption, increased vaping, and stronger psychological dependence on smoking (26).

Five studies examined the relationship between social isolation and smoking behaviors [6,8,21, 32, 33]. Findings suggest that social isolation was associated with less favorable smoking outcomes. Compared with non-smokers or never smokers, current smokers reported lower levels of social and emotional support and were more likely to experience social isolation. Individuals living with others were more likely to reduce smoking and report intentions to quit, suggesting that household support may facilitate smoking reduction [32]. Similarly, smoking abstinence was associated with transitions toward larger social networks with lower smoking exposure, indicating that cessation may strengthen social connectedness while reducing exposure to smoking (6, 8, 21, 32, 33).

#### COVID-19 impacts on loneliness, isolation, and smoking behaviors

Six studies outside the meta-analysis examined the impact of the COVID-19 pandemic and related social restrictions on smoking behaviors (6, 8, 25, 26, 29, 30). Findings suggested that pandemic-related social disruption influenced smoking behaviors, although the direction of these associations varied across populations. Three studies reported statistically significant associations between greater loneliness or social disconnection during the pandemic and increased smoking or vaping. One study specifically reported that cigarette smokers had significantly lower levels of social and emotional support than never smokers and former smokers [7].

Two studies found evidence of reduced socially motivated smoking during periods of restricted social interaction [25, 31]. Reduced interpersonal contact during lockdown was associated with less social smoking and cigarette sharing [31], and smoking abstinence was associated with transitions toward larger social networks with lower smoking exposure [25]. In contrast, one study reported a statistically significant inverse association, finding that greater loneliness was associated with lower odds of smoking among older adults, with no significant association between reduced in-person contact and smoking [27]. Collectively, these findings suggest that the COVID-19 pandemic influenced smoking behaviors through changes in social connectedness, with most studies indicating that loneliness and social isolation increased nicotine product use.

#### Psychological Distress, Mental Health, and Smoking

Four studies outside the meta-analysis examined psychological distress, emotional coping, or mental health factors in relation to loneliness, isolation, and smoking behaviors [24, 27, 28, 36]. Overall, findings suggest that smoking may function as a coping mechanism for psychological distress and perceived social disconnection. Three studies reported positive associations between psychological distress or loneliness-related coping and smoking behaviors, indicating that greater loneliness or distress was associated with heavier cigarette consumption, stronger psychological dependence, and smoking as a strategy for coping with isolation and stress [24, 27, 36].

One study reported mixed findings, indicating that smoking and vaping behaviors were significantly associated with loneliness within participants’ social networks, but not with participants’ own self-reported loneliness. Collectively, these findings suggest that emotional distress, loneliness, and perceived social disconnection may contribute to smoking initiation, maintenance, and dependence [28].

## Discussion

This systematic review and meta-analysis synthesized published literature between 2015 and 2025 to examine the relationship between loneliness, or social isolation, and nicotine use behaviors in adults. The study’s findings indicate a consistent and statistically significant association of social isolation or loneliness on nicotine product use. The meta-analysis indicated that social isolation or loneliness was associated with an 84% increase in the odds of nicotine product use compared with non-isolated individuals, although substantial between-study heterogeneity was observed. Importantly, this heterogeneity across studies in the meta-analysis was very high (I^2^=92.3%), indicating substantial variability in effect sizes across studies. In accounting for this high heterogeneity, leave-one-out sensitivity analysis demonstrated that the pooled effect remained stable and statistically significant regardless of which study was omitted. Overall, these findings contribute to the growing evidence base supporting the importance of social isolation and loneliness as an important psychosocial determinants of nicotine product use. In contrast, the second meta-analysis, which examined the reverse direction of this relationship (the impact of nicotine product use on subsequent social isolation or loneliness) found no statistically significant association (OR = 1.35, 95% CI: 0.72–2.53), with similarly substantial heterogeneity (I² = 92.9%). However, this analysis was based on only two studies, yielding three effect estimates, and should therefore be interpreted with caution given the limited evidence base.

This study builds on a previous systematic review, conducted by Dyal and Valente (16), which concluded that loneliness and smoking were associated, but that effect sizes were relatively small. Our findings demonstrate a substantially larger and methodologically diverse body of literature published in the subsequent decade, particularly during periods of sustained social isolation, such as the COVID-19 pandemic. The pooled effect observed in the present review suggests a stronger association between social isolation or loneliness and nicotine product use than previously reported in the Dyal and Valente review (16). Findings from the studies conducted during the COVID-19 pandemic suggest that loneliness and social isolation were more strongly associated with nicotine product use during lockdown. The narrative analysis found that increased loneliness, social isolation, and distress were associated with increased nicotine product use, potentially resulting from increased stress, a demonstrated influence on nicotine product use behaviors (43, 44).

Subgroup analyses revealed a moderately stronger association with nicotine product use for studies measuring social isolation compared with those measuring loneliness, suggesting that objective indicators of social isolation may exert a greater influence on nicotine use than subjective loneliness. It is important to note that social isolation and loneliness were not consistently separated in all subgroup analyses, which limited direct comparability across studies. To address this heterogeneity, a meta-regression was conducted, finding no statistically significant difference between the two constructs, suggesting that the apparent divergence in subgroup estimates should be interpreted with caution. This finding merits further exploration and addressing in both nicotine and other substance use research. Several mechanisms have been proposed to explain these associations. Loneliness and social isolation may contribute to nicotine use through psychosocial pathways, including increased stress, poorer coping capacity, and reduced access to social support (2, 3). Individuals experiencing loneliness may engage in nicotine use as a maladaptive coping strategy to alleviate negative affect, while limited social networks may reduce exposure to health-promoting norms and decrease opportunities for cessation support (2, 3).

In conducting sub-group analysis, social isolation or loneliness had a stronger association with e-cigarette use than combustible cigarette smoking although results should be interpreted cautiously as the vaping subgroup was based on a very limited number of studies (n=2) (22, 31) and meta-regression analysis showed no statistically significant differences between them. The observed relationship between social isolation or loneliness and nicotine product use was found to be robust across different outcome definitions, though the persistently high heterogeneity across both subgroups points to other unmeasured sources of variability, such as population characteristics or study design, rather than the choice of nicotine use measure itself.

Age group-based subgroup analyses also demonstrated important findings, with middle aged (age 26-40 years) and upper and middle-aged (age 41-60 years) individuals demonstrating stronger associations between social isolation or loneliness and nicotine product compared to seniors (age 61+ years), consistent with prior literature (45). This finding suggests that social disconnection may have a particularly important influence on nicotine use during age periods characterized by social role, employment, and caregiving transitions, warranting additional research. These findings complement the narrative analysis results by indicating that social and peer-related influences exert a comparatively stronger effect on smoking behaviors among younger populations (as observed in adolescents) (46) whereas smoking in older adults may be maintained through chronic dependence and psychosocial vulnerability, as seen in the literature (47). Therefore, while social isolation may influence socially motivated smoking reductions, distress-induced smoking influenced by isolation may increase nicotine product use (48). Across the studies examined, psychological distress, particularly loneliness, anxiety, depression, and emotional distress, frequently co-occurred with smoking (29, 32, 33, 37, 39), consistent with established links between smoking and affect regulation (49, 50). Psychological distress, perhaps resulting from social isolation, may lead to smoking as a coping strategy. This has been demonstrated in recent findings which position distress as a bridging relationship between smoking and loneliness (51). Further research should explore this relationship.

## Limitations

Limitations specific to this review include the restriction to studies published in English, which may have resulted in the exclusion of relevant research published in other languages and global regions. This review did not employ a formal certainty-of-evidence framework such as GRADE, which limits the ability to make standardized statements about confidence in the pooled estimates. Additionally, the heterogeneity of the studies was consistently high across the overall meta-analysis and subgroup analyses. This may demonstrate high levels of diversity amongst studies based on populations, measurements, and findings. Consequently, results should be interpreted with caution, as meta-analysis and subgroup analyses may mask true effects, or in some cases invalidate pooled estimates. Studies were combined if reporting ORs and RRs as effect estimates in the meta-analysis, which may raise concerns about the validity of the pooled estimate. However, sensitivity analyses conducted separately for studies reporting ORs and RRs yielded consistent results, with both groups demonstrating an increased likelihood of nicotine product use among individuals experiencing social isolation or loneliness. Most included studies were cross-sectional in design (32, 34, 37, 41), limiting the ability to establish causal or temporal relationships between loneliness, social isolation, and nicotine product use. Another limitation is the differences between studies in their measurement of loneliness and social isolation. A wide range of instruments was reported, including single-item loneliness measures, composite social isolation indices, and validated scales such as the UCLA Loneliness Scale and ENRICHD Social Support Instrument, affecting comparability across studies. This variability may limit comparability across studies and complicate the synthesis. Moreover, many studies relied on self-reported smoking and psychosocial measures, which may be subject to recall bias and social desirability bias.

### Practice & Research Implications

Important gaps in the literature were also identified. Few studies examined racialized populations, 2SLGBTQ+ communities, or other marginalized groups disproportionately affected by both nicotine use and social isolation. Similarly, limited evidence explored sex- and gender-specific mechanisms, intersectionality, or socioeconomic pathways linking loneliness and smoking. Longitudinal and intervention studies are also needed to clarify mediating mechanisms, including psychological distress and social support. Nicotine use cessation strategies may benefit from integrating mental health screening, emotional support, and social connectedness interventions alongside traditional nicotine dependence treatments, particularly for vulnerable populations such as older adults, socially isolated individuals, and those experiencing psychological distress. Additional research should examine the effect of social isolation and nicotine product use on different age groups to elucidate the nuanced factors that may be involved in the older adults having a weaker effect than middle aged groups, such as differing quit motivations by age group (52) or availability and acceptability of age specific programming (53). As social isolation and loneliness were not found to have significantly different effects on nicotine product use in the meta-regression, these concepts may be more closely related than distinct. However, further research that differentiates these two concepts as distinct, though related, should be embedded in study design, measurement tools, and reporting practices to further elucidate the effects of objective lack of social contact with subjective experience of loneliness. Future research should adopt consistent methodology, definitions, and measurement scales to enable clearer comparison across studies.

## Conclusion

Overall, the evidence suggests that loneliness and social isolation are associated with nicotine product use across diverse populations. Tentative evidence points to a bidirectional relationship between social isolation, loneliness, and nicotine product use, suggesting that these factors are interrelated. Psychological distress and emotional vulnerability further appeared to strengthen these relationships, particularly during the COVID-19 lock down. These findings suggest that stronger social support and social connectedness may support nicotine product use reduction and cessation. Future research should prioritize longitudinal and intervention research to better clarify causal pathways and mediating factors and to inform smoking cessation strategies.

## Data availability

Data will be made available upon request to the corresponding author.

## Supplementary Material

**Supplementary Figure 1.**
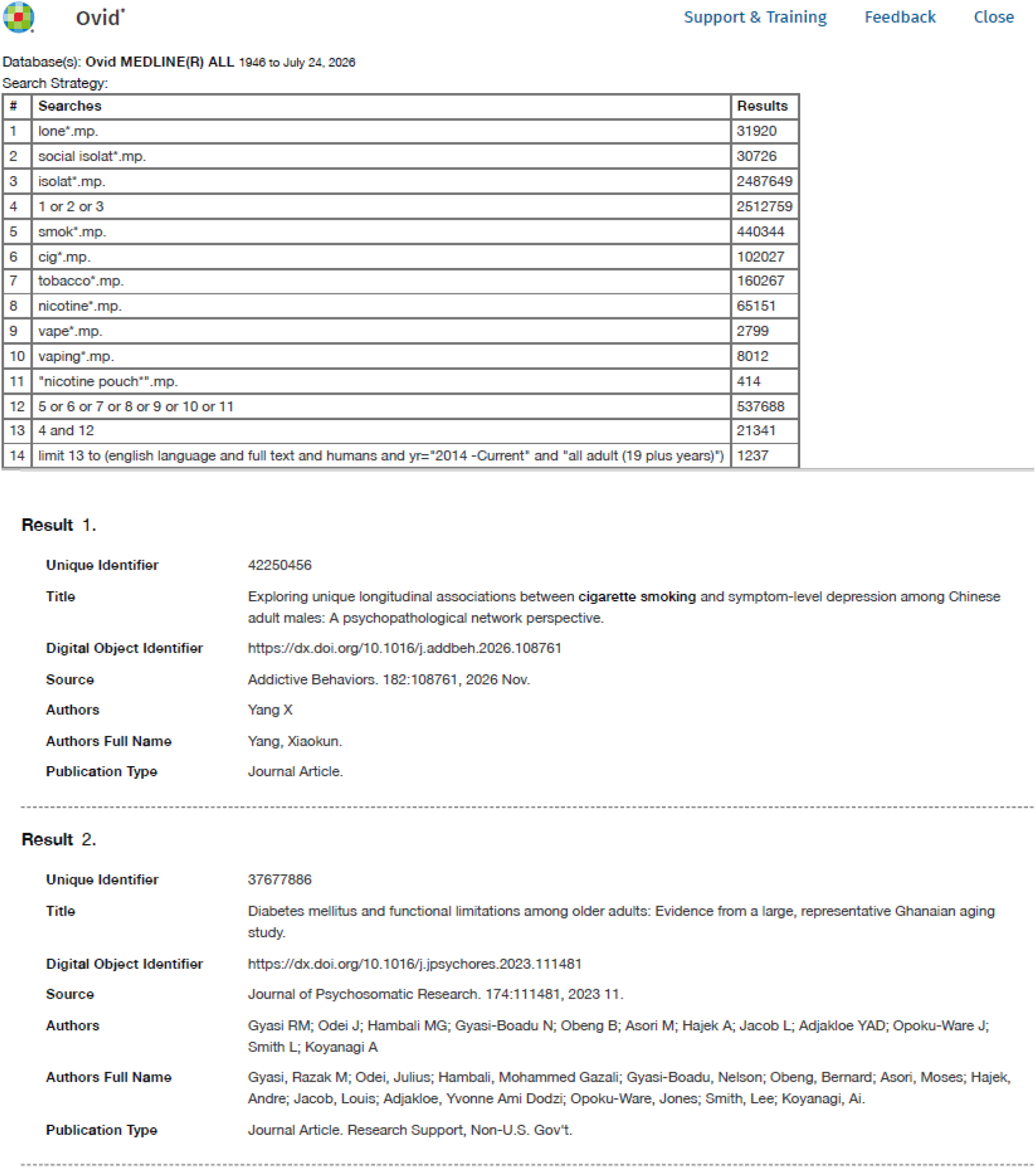
MEDLINE Search Strategy.

**Supplementary Figure 2.**
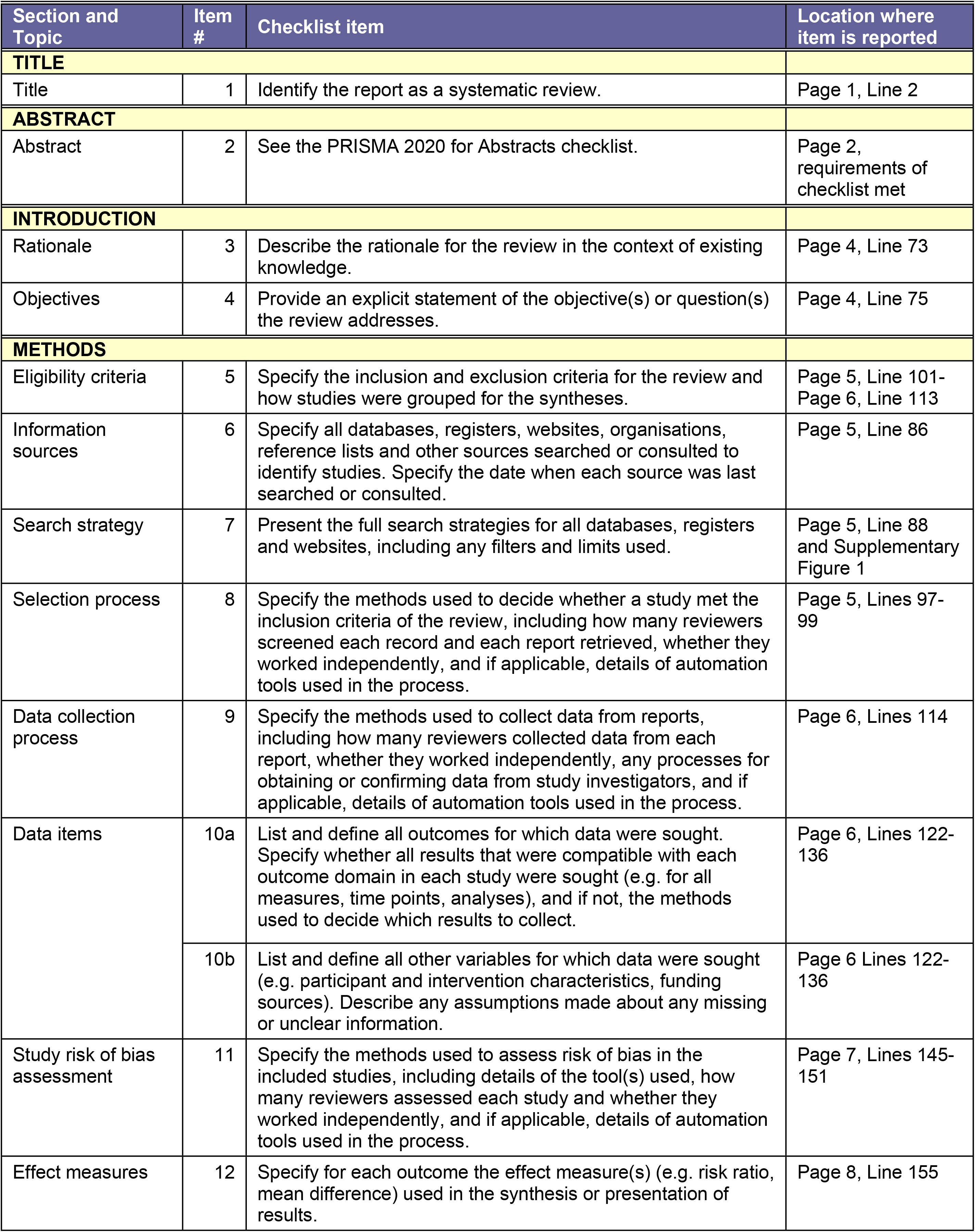
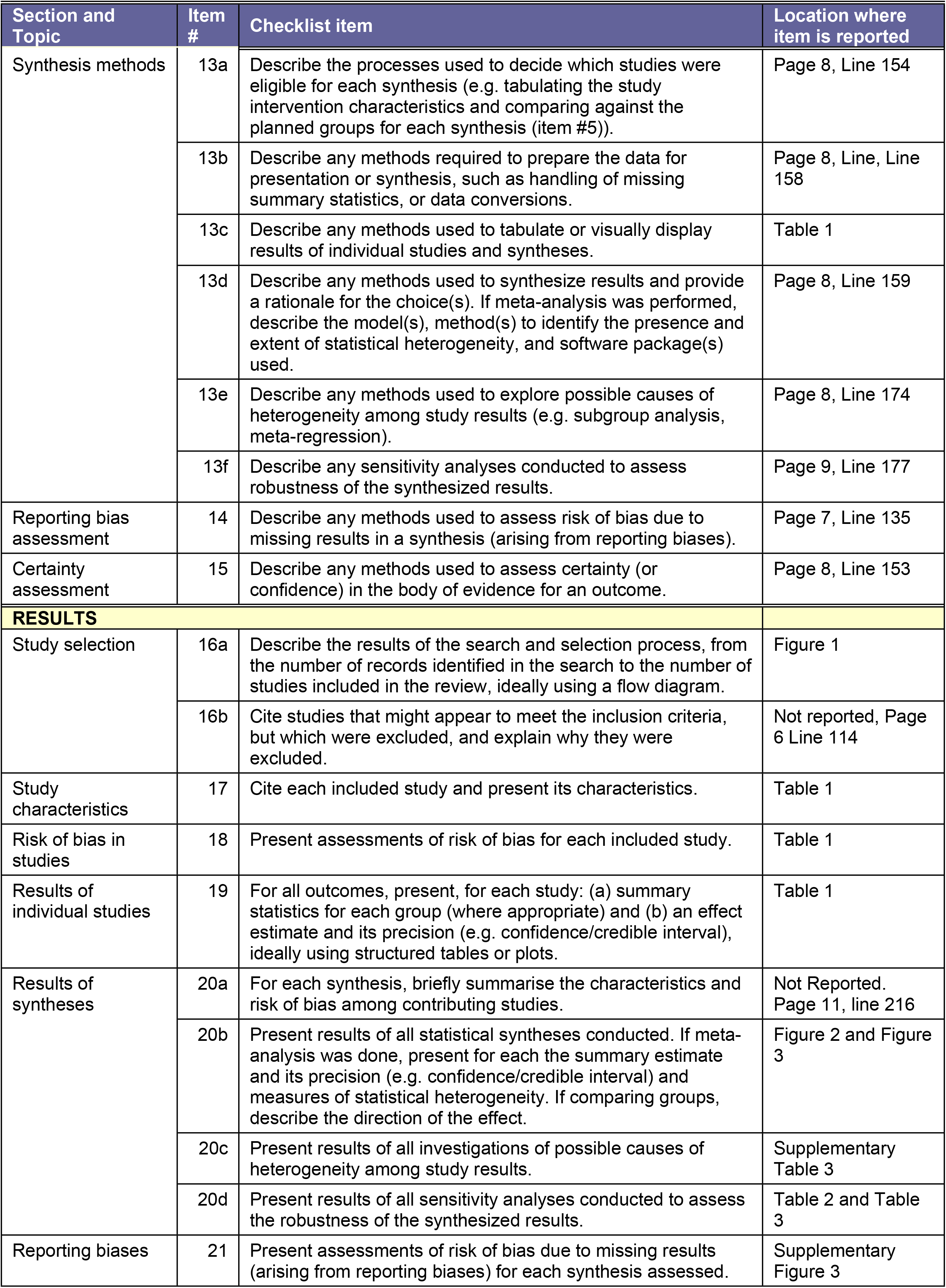
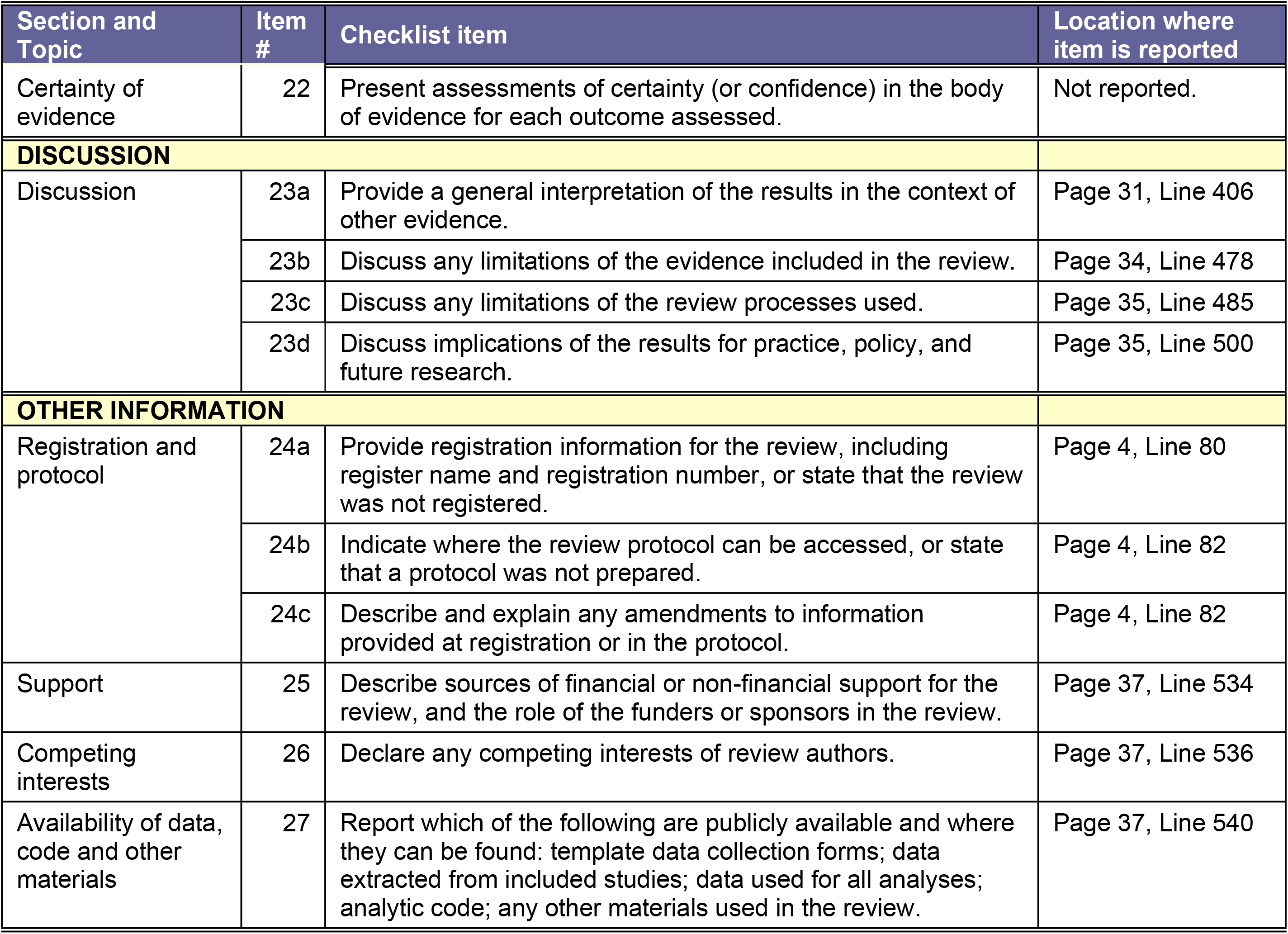
PRISMA Reporting Guidelines.

**Supplementary Figure 3.**
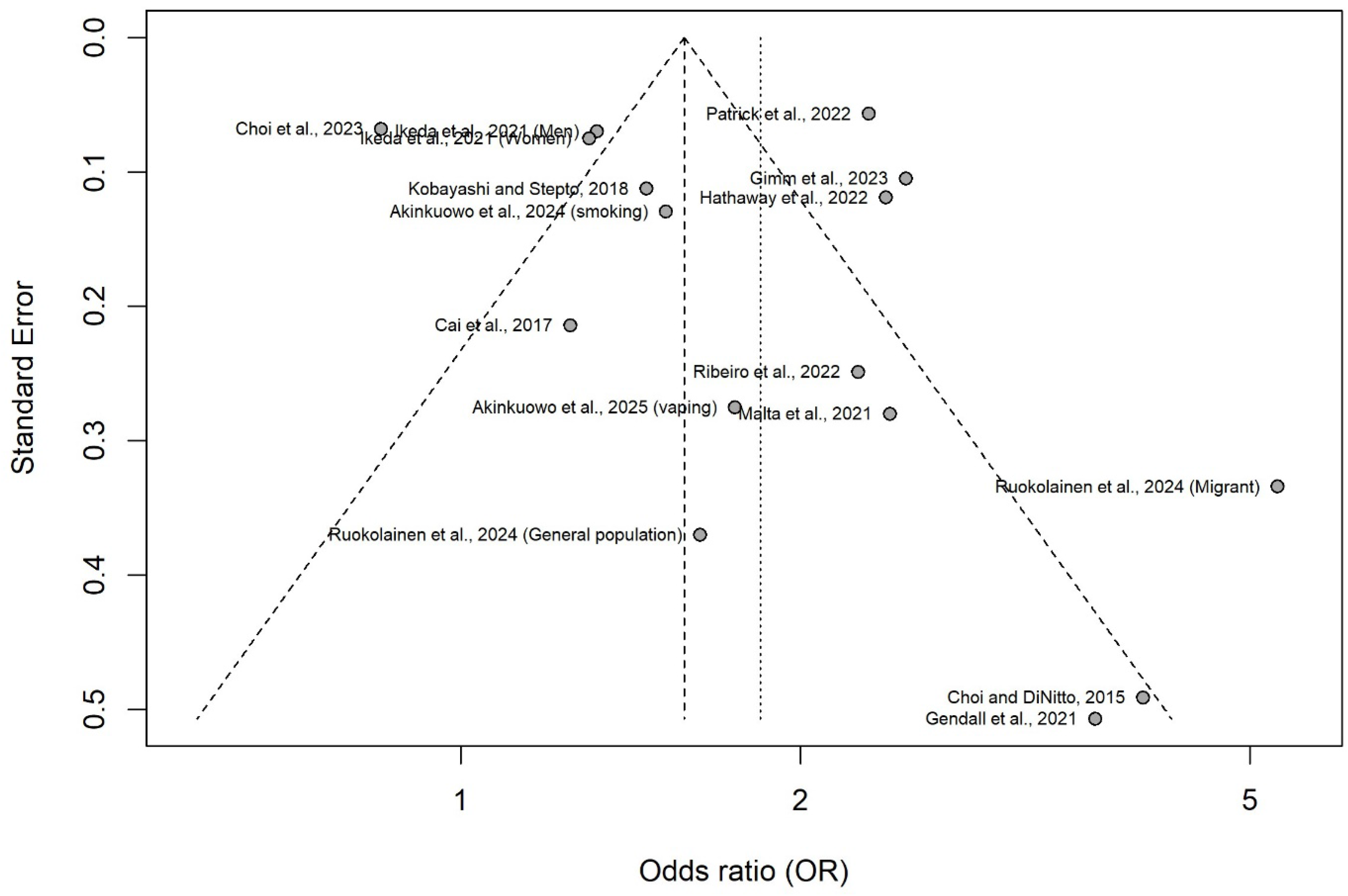
Forest Plot of Overall Effects.

**Supplementary Figure 4.**
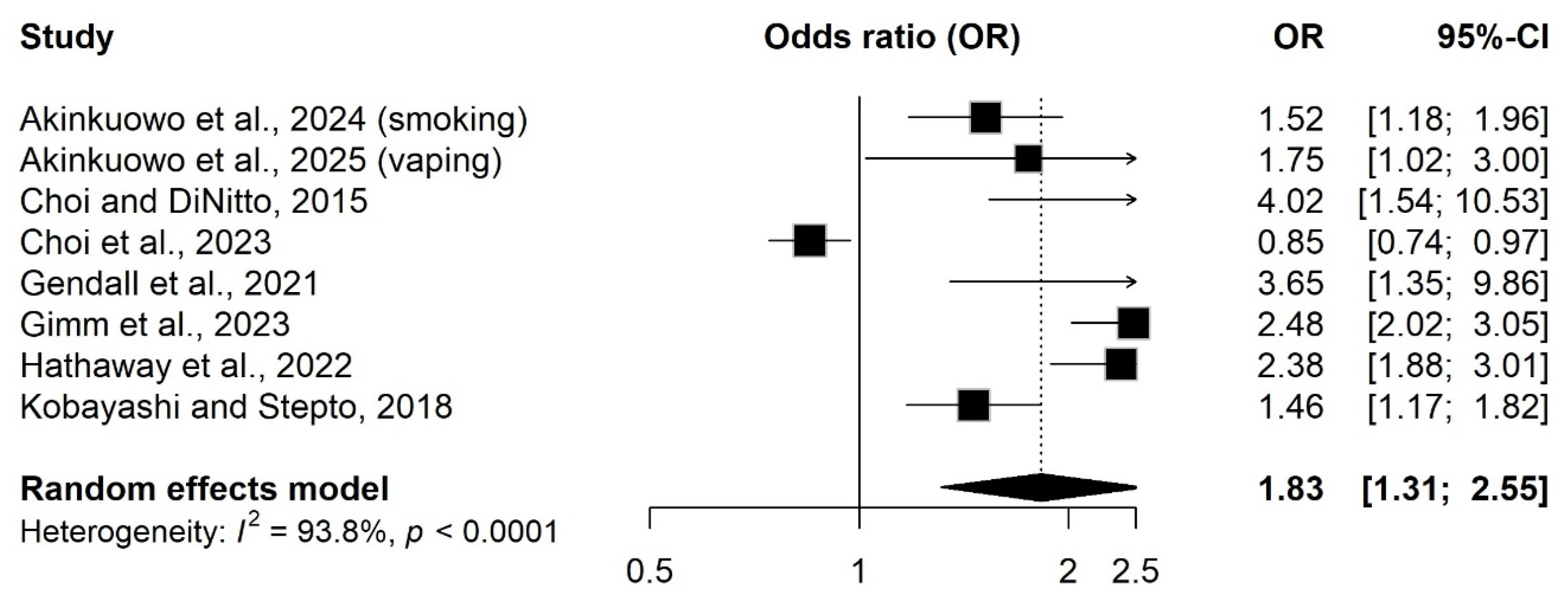
Forest Plot of Studies Reporting OR only.

**Supplementary Figure 5.**
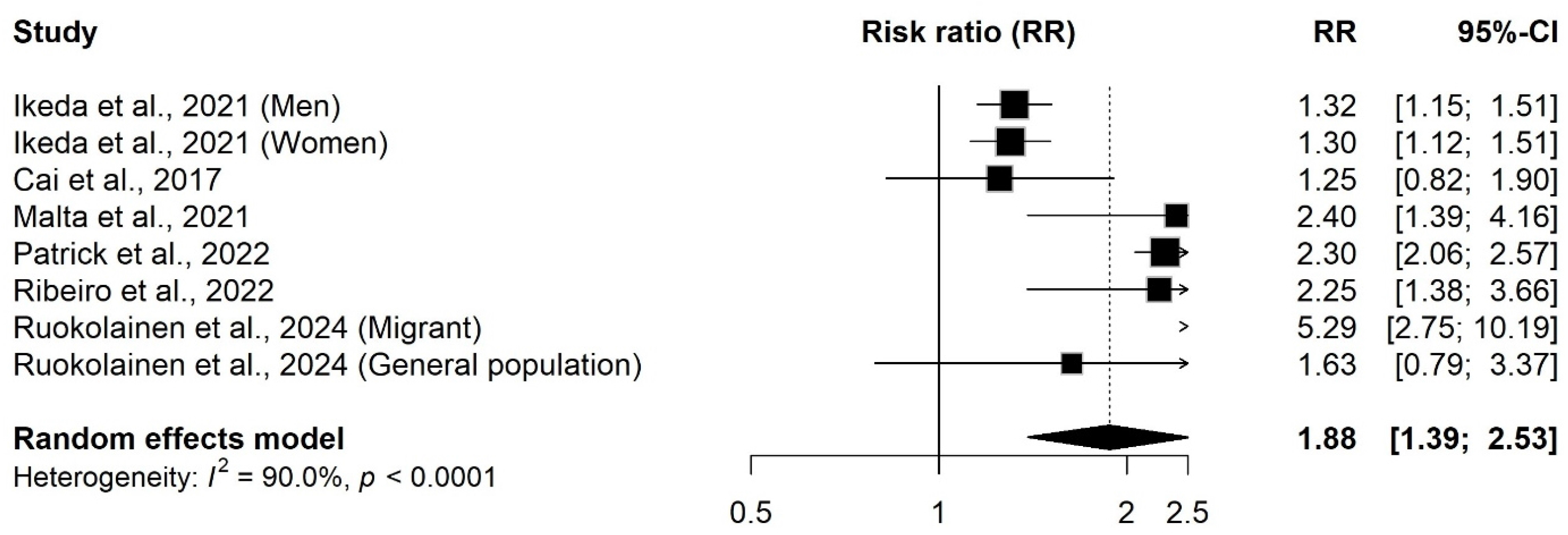
Forest Plot of Studies Reporting RR online.

**Supplementary Table 3.** Meta-regression examining statistical significance between subgroups.

| ***Moderator Variable*** | ***Reference*** | ***Co-efficient (95% CI)*** | ***Q-test for moderators:***<br>***QM (df)*** | ***P-value*** |
| --- | --- | --- | --- | --- |
| ***Exposure type*** |  |  | 0.70 (2) | 0.705 |
| *Social isolation* | *Loneliness* | 0.20 (-0.28 – 0.68) |  |  |
| *Both outcomes* | *Loneliness* | 0.15 (-0.63 – 0.92) |  |  |
| ***Product type*** |  |  | 0.14 (1) | 0.711 |
| *E-cigarette* | *Cigarette* | 0.12 (-0.53 – 0.78) |  |  |
| ***Age group*** |  |  | 3.27 (3) | 0.352 |
| *Middle age (26-40)* | *Young adult (18-25)* | 0.45 (-0.36 – 1.27) |  |  |
| *Upper middle age (41-60)* | *Young adult (18-25)* | 0.20 (-0.57 – 0.96) |  |  |
| *Senior (61+)* | *Young adult (18-25)* | -0.11 (-0.76 – 0.55) |  |  |
| ***Measures of nicotine product use*** |  |  | 0.11 (1) | 0.741 |
| *Current use* | *Increased use* | -0.08 (-0.53 – 0.38) |  |  |
| ***COVID-19*** |  |  | 0.44 (1) | 0.507 |
| *Before COVID-19* | *After COVID-19* | -0.15 (-0.60 – 0.30) |  |  |

**Supplementary Figure 6.**
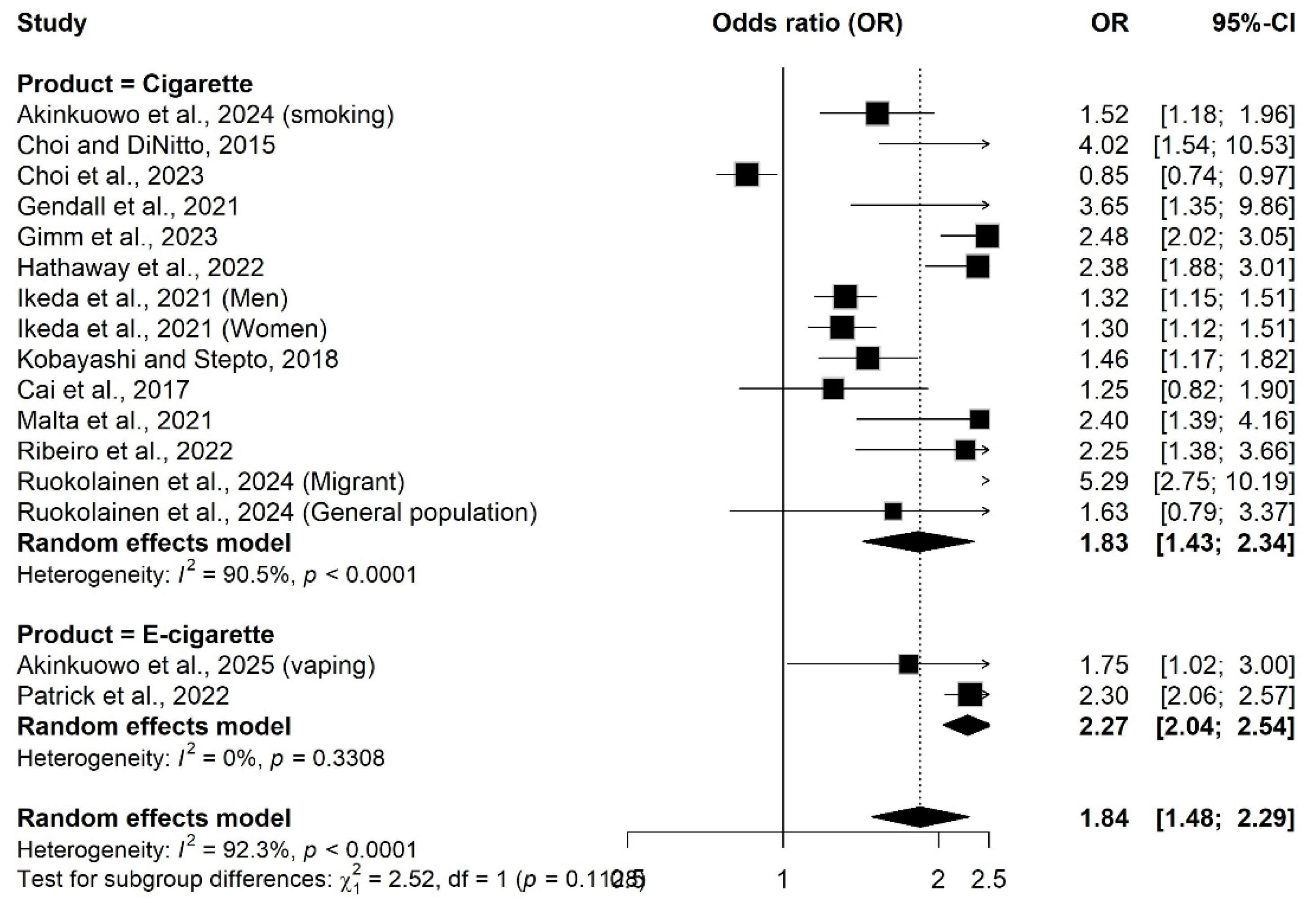
Subgroup analysis showing effects of social isolation and loneliness on cigarettes compared to e-cigarette use.

**Supplementary Figure 7.**
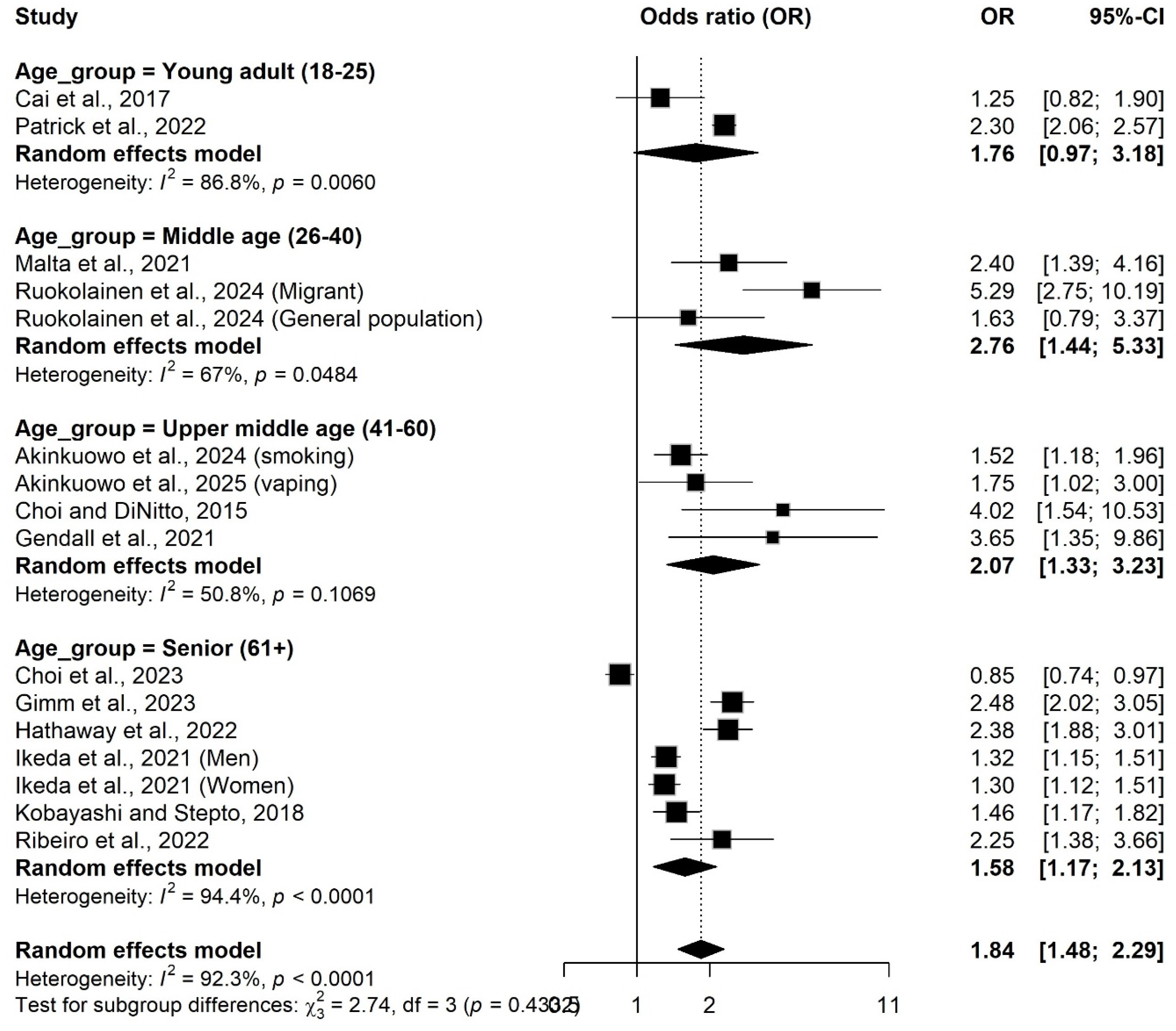
Subgroup analysis showing effects of social isolation and loneliness on nicotine product use for different age categories.

**Supplementary Figure 8.**
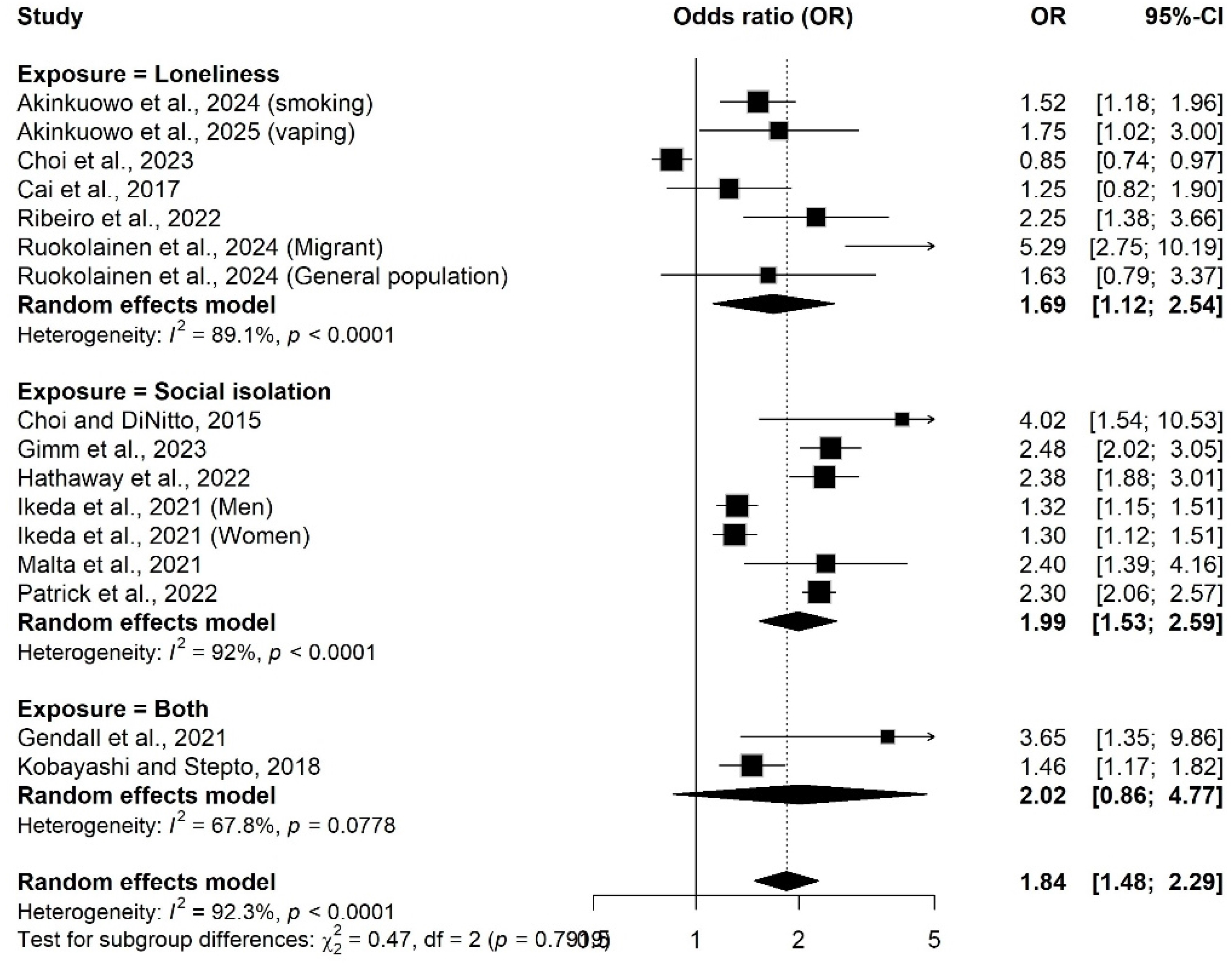
Subgroup analysis showing effects of social isolation compared to loneliness on nicotine product use.

**Supplementary Figure 9.**
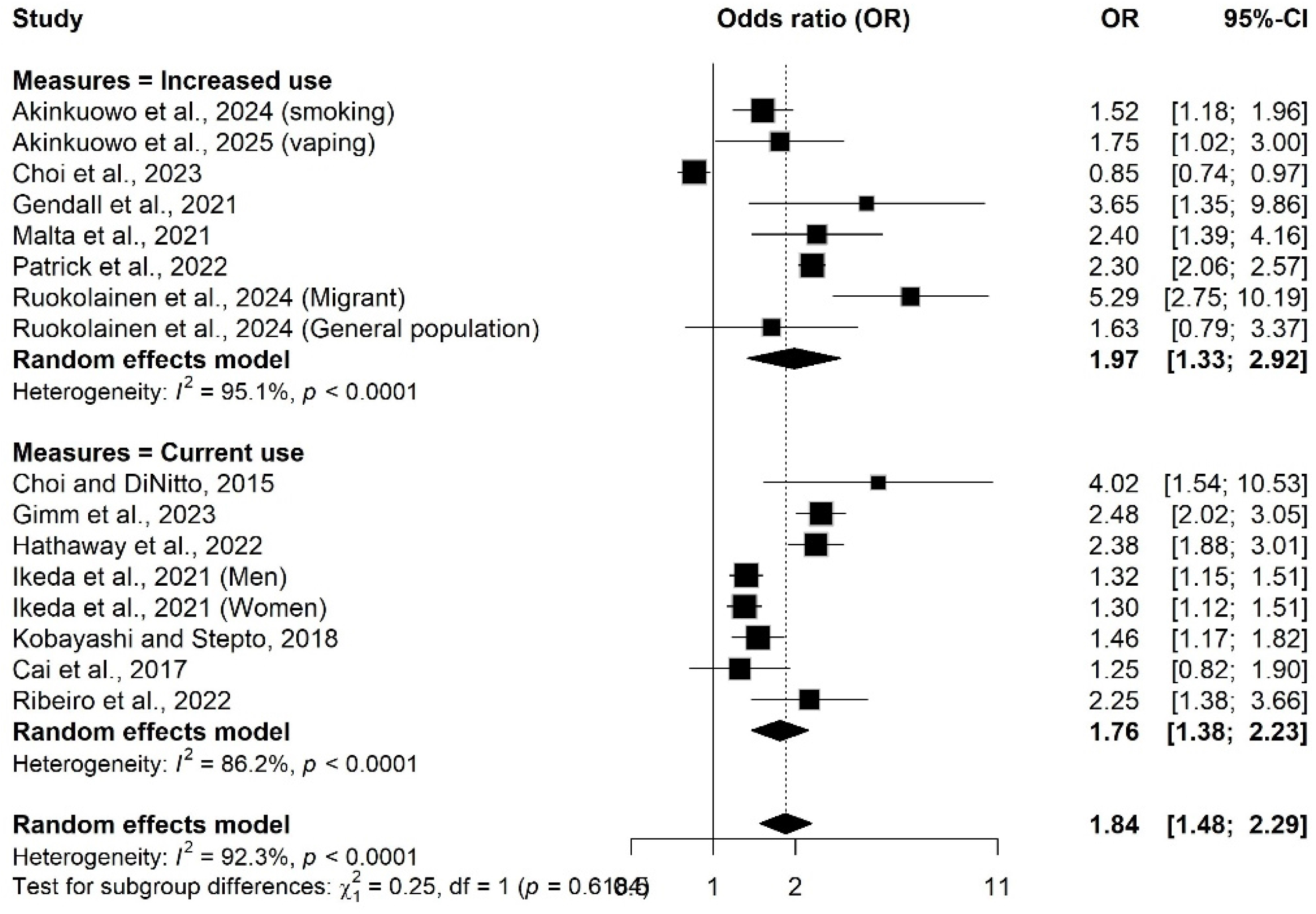
Subgroup analysis showing effects of social isolation and loneliness on nicotine product use based on measures of nicotine use behaviour.

**Supplementary Figure 10.**
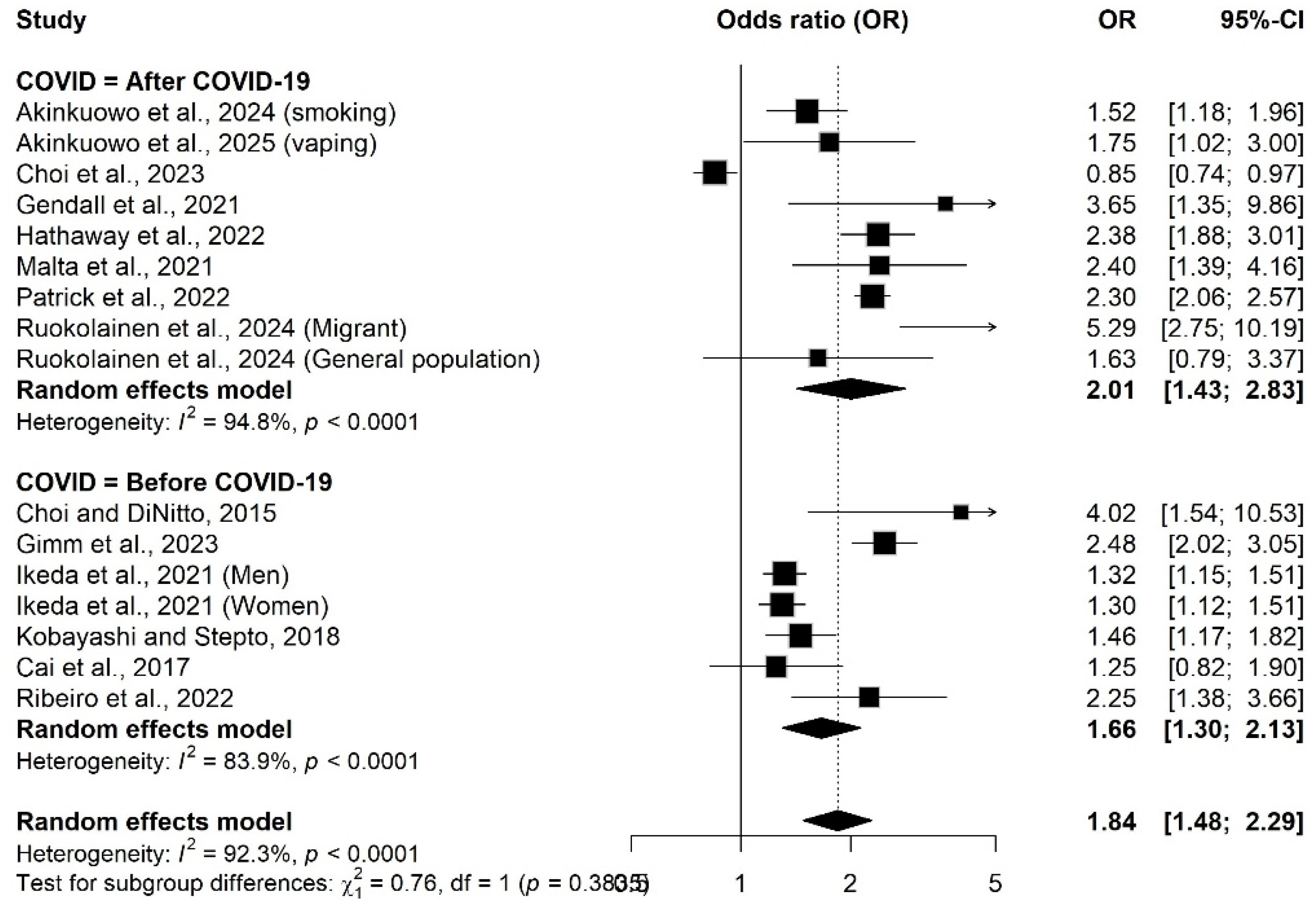
Subgroup analysis showing effects of social isolation and loneliness after COVID-19 on nicotine product use compared to before COVID-19.

